# Electromechanical Drivers of Diastolic Dysfunction During Hypertrophic Cardiomyopathy Progression

**DOI:** 10.64898/2026.11.10.26367717

**Authors:** Abdallah I Hasaballa, James A Coleman, Zhinuo J Wang, Ruben Doste, Xin Zhou, Alberto Zingaro, Mariano Vázquez, Alessia Argirò, Raffaele Coppini, Iacopo Olivotto, Alfonso Bueno-Orovio

**Author notes:** Corresponding author: Alfonso Bueno-Orovio.

## Abstract

**Aims:** Diastolic dysfunction in hypertrophic cardiomyopathy (HCM) reflects changes in sarcomeric function, electrophysiology, ventricular structure, and myocardial mechanics. Their effects on filling remain difficult to determine clinically. We sought to dissect these effects in MYH7-related non-obstructive HCM progression.

**Methods:** We developed a human biventricular electromechanical model linking electrophysiology and calcium handling to contraction, ventricular mechanics, haemodynamics, electrocardiography, and myocardial strain. Stage I represented MYH7 sarcomeric dysfunction before hypertrophy; Stage II added septal hypertrophy; Stage III added increased myocardial stiffness. Secondary ionic remodelling was examined at each stage and severity.

**Results:** Sarcomeric dysfunction alone generated a low-volume hypercontractile phenotype before overt hypertrophy. Septal hypertrophy further constrained chamber filling, whereas myocardial stiffening exacerbated diastolic dysfunction and regional mechanical heterogeneity. Secondary ionic remodelling produced marked repolarisation abnormalities but, importantly, also prolonged calcium–tension dynamics and impaired relaxation, reducing ventricular filling to an extent comparable with structural remodelling in some disease states. Increasing ionic-remodelling severity progressively amplified these effects. Despite preserved ejection fraction (EF) throughout progression, ventricular volumes decreased while global and regional myocardial deformation deteriorated.

**Conclusion:** Diastolic dysfunction was evident before overt hypertrophy and worsened as structural, mechanical, and ionic abnormalities were added. Preserved EF did not reflect reduced ventricular filling or deterioration in myocardial strain. Regional mechanical measures may therefore reveal disease burden not captured by EF alone. By separating individual disease mechanisms, in silico modelling may help link measurable biomarkers to their underlying causes and support mechanism-based patient stratification.

Graphical Abstract
Translational Perspective Preserved ejection fraction may obscure clinically relevant disease burden in hypertrophic cardiomyopathy, as impaired filling appears to arise from more than hypertrophy alone. Our staged human biventricular electromechanical simulations suggest that sarcomeric dysfunction, septal hypertrophy, increased myocardial stiffness, and secondary ionic remodelling make distinct, yet interacting, contributions to diastolic impairment. Regional strain and active-stress measures may therefore reveal abnormalities that are not apparent from ejection fraction, while ECG and pressure–volume indices provide complementary information. Together, these markers could help distinguish underlying disease mechanisms and support more mechanistically informed patient stratification.

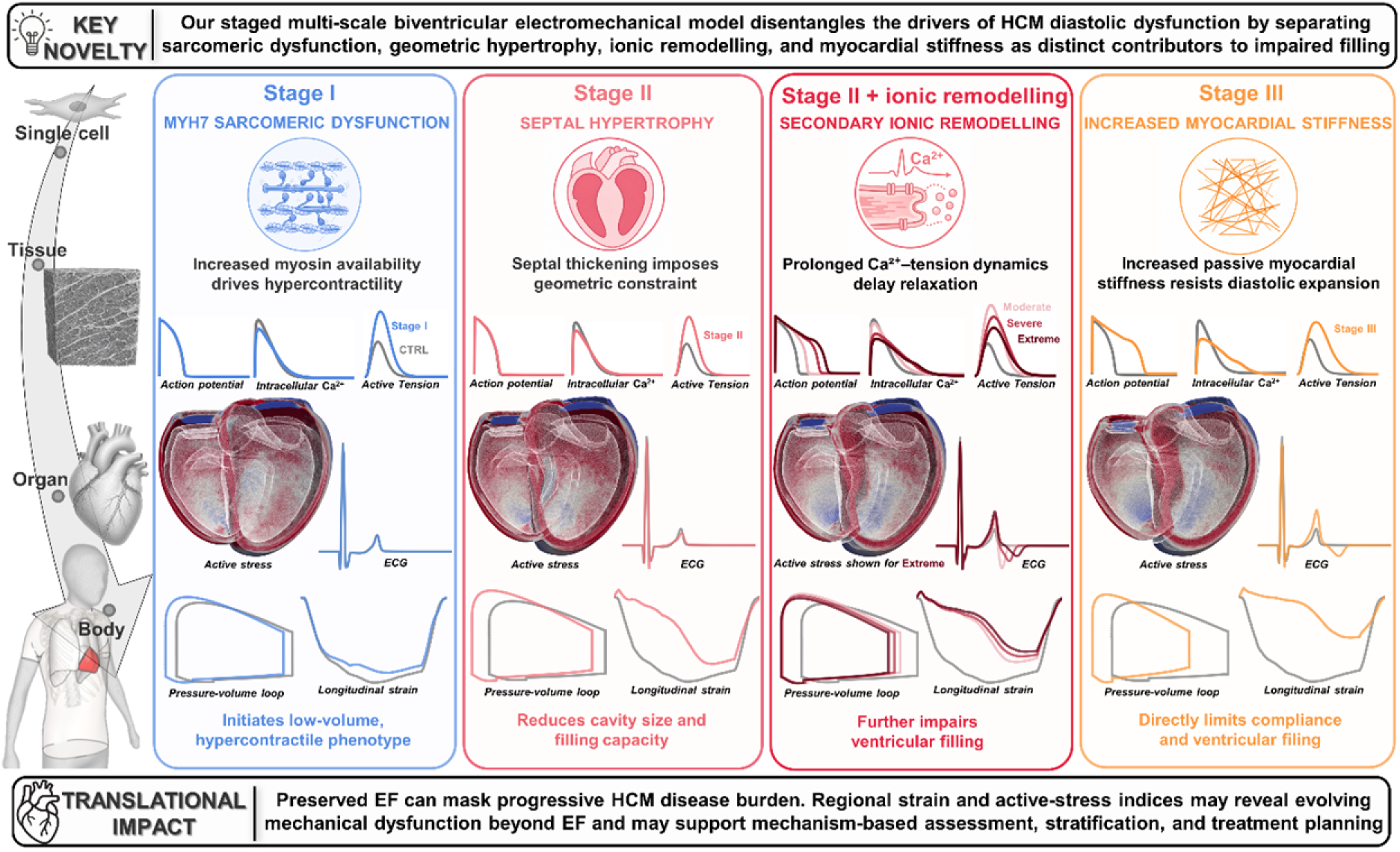

## 1. Introduction

Hypertrophic cardiomyopathy (HCM) is the most common inherited myocardial disease and remains a major cause of heart failure, arrhythmia, and sudden cardiac death in the young. Although clinically defined by otherwise unexplained left ventricular (LV) hypertrophy, HCM is a heterogeneous disease rather than a single phenotype, ranging from genotype-positive individuals without overt hypertrophy to advanced myocardial remodelling with impaired filling, arrhythmic risk, and progressive heart failure^1–4^. This variability reflects not only genetic diversity, but also the interaction between sarcomeric dysfunction, electrophysiological remodelling, altered calcium handling, myocardial hypertrophy, and increased ventricular stiffness. Consequently, defining how these mechanisms contribute to clinically measurable phenotypes across different stages of disease remains a major challenge in HCM. This challenge is particularly pronounced in non-obstructive HCM, where the driving pathophysiology remains incompletely understood, and therapeutic options are more limited than in obstructive disease^5^.

Most familial HCM is caused by pathogenic variants in genes encoding sarcomeric proteins, which alter force generation, relaxation, and myocardial energetics^6,7^. Among these, MYH7, encoding β-myosin heavy chain, is one of the most frequently implicated genes and is central to thick-filament regulation. Experimental studies have shown that HCM-associated myosin variants can destabilise energy-conserving myosin states, increase myosin head availability for cross-bridge formation, and promote hypercontractility with impaired relaxation^8^. However, sarcomeric dysfunction alone does not fully account for the organ-level HCM phenotype. Secondary ionic remodelling, altered calcium and sodium homeostasis, reduced repolarisation reserve, anatomical hypertrophy, and increased myocardial stiffness all contribute to the evolving electromechanical phenotype^9,10^. How these mechanisms interact across scales and contribute differentially across disease stages remains incompletely understood.

Clinical imaging, electrocardiography (ECG), and haemodynamic assessment provide essential markers of HCM severity, but they cannot readily isolate causal mechanisms. Pressure–volume indices quantify chamber-level performance, surface ECGs capture activation and repolarisation abnormalities, and strain imaging can reveal myocardial dysfunction before global ejection fraction (EF) is reduced^11,12^. Yet, these measurements represent integrated outputs of multiple interacting processes. Consequently, it remains difficult to determine how mutation-driven myofilament dysfunction, secondary electrophysiological remodelling, hypertrophy, and myocardial stiffening individually and collectively shape disease severity and adverse remodelling.

Human multiscale cardiac modelling and simulation offers a mechanistic framework to address this gap. Previous computational studies in HCM have provided insight into electrophysiological substrates and arrhythmic risk, but have largely focused on electrical rather than coupled electromechanical phenotypes^13,14^. By integrating cellular electrophysiology, calcium handling, active tension generation, ventricular mechanics, circulatory loading, pressure–volume dynamics, ECG, and myocardial deformation within a controlled *in silico* environment, multiscale electromechanical models can link disease mechanisms to clinically measurable biomarkers^15–25^. This approach enables individual disease components to be perturbed systematically, allowing the primary effects of sarcomeric dysfunction to be separated from secondary ionic, geometric, and mechanical remodelling. In HCM, such a framework is particularly suited to investigating how myofilament abnormalities and downstream remodelling shape ventricular haemodynamics, ECG morphology, myocardial strain, and regional stress generation.

In this study, we used a human biventricular electromechanical modelling framework to investigate MYH7-related non-obstructive HCM across disease states of increasing severity. To our knowledge, this is the first three-dimensional (3D) human biventricular study to combine MYH7-related sarcomeric dysfunction with graded regional ionic remodelling, septal hypertrophy, and increased myocardial stiffness at the organ scale. Starting from a calibrated healthy model, these components were introduced sequentially to separate their individual and combined effects on electrical, haemodynamic, and mechanical phenotypes. Secondary ionic remodelling was represented by graded HCM-associated changes in ionic currents and calcium handling derived from human septal myectomy data. We hypothesised that sarcomeric dysfunction would produce an early low-volume hypercontractile phenotype, while hypertrophy and stiffness would progressively restrict filling, and ionic remodelling would prolong repolarisation and calcium-driven tension, impair relaxation, and further compromise filling. This framework therefore provides a mechanistic basis for understanding the processes that drive HCM phenotype expression and the emergence of clinically relevant biomarkers of adverse remodelling.

## 2. Methods

### 2.1. Multiscale human biventricular modelling and simulation framework

A multiscale, multiphysics, human biventricular electromechanical modelling framework was developed to link cellular disease mechanisms with organ-scale electrical, haemodynamic, and mechanical phenotypes. It was used to construct a calibrated healthy-control model and staged HCM phenotypes, enabling systematic comparison of the contributions of individual disease mechanisms across stages. The modelling approach builds on previously established human ventricular electromechanical models^20–24^. Additional methodological details are provided in the Supplementary Material. Electromechanical simulations were performed using Alya, a highly parallel multiphysics simulation code developed at the Barcelona Supercomputing Center^26,27^.

#### 2.1.1. Anatomical model

A biventricular finite-element mesh composed of linear tetrahedral elements, with the valvular openings closed to form enclosed ventricular cavities, was derived from four-chamber cardiac computed tomography geometries of a healthy human heart obtained from a published database^28^. The mesh had a mean equivalent tetrahedral edge length of 1.02 mm. Rule-based fibre, sheetlet, and sheet-normal orientations were assigned throughout the myocardium to define orthotropic electrical conduction and mechanical behaviour^29^.

#### 2.1.2. Electrophysiological model

Electrical conduction was simulated using a monodomain formulation with orthotropic conduction aligned to myocardial microstructure. Cellular electrophysiology and calcium handling were represented using the human ventricular ToR-ORd model^30^, incorporating physiological transmural and apico-basal heterogeneity^31^. Ventricular activation was initiated through an endocardial fast-activation layer mimicking the trifascicular conduction system and Purkinje–myocardial junctions^32^. The resulting activation sequence was calibrated against healthy 12-lead ECG biomarkers using an established ECG-informed approach^15^. Pseudo-ECGs were computed at standard 12-lead electrode locations.

#### 2.1.3. Mechanical model

Ventricular mechanics were modelled using a 3D dynamic hyperelastic active stress total-Lagrangian finite-element formulation. Passive myocardial behaviour was represented using a nearly incompressible Holzapfel–Ogden constitutive law^33,34^, with physiological boundary conditions applied to reproduce ventricular loading, ejection, and pericardial restraint. Ventricular outflow was coupled to reduced-order systemic and pulmonary circulations through 2-element Windkessel models, enabling simulation of pressure–volume dynamics under physiological loading conditions.

#### 2.1.4. Electromechanical coupling

Electromechanical coupling followed the previously established bidirectionally coupled framework^20,22^ linking ToR-ORd calcium transients to active tension generation through the Land myofilament formulation^35^. Intracellular calcium dynamics were coupled to calcium binding to troponin, with troponin-bound calcium contributing to intracellular calcium buffering, while myocardial stretch and stretch rate influenced active-tension development. Mechanical deformation also influenced electrical propagation through deformation-dependent diffusion. Parameters of the original Land model had been re-calibrated previously^22^ to achieve physiological active tension peaks when driven by ToR-ORd calcium transients. The resulting active stress was added to the passive myocardial stress tensor in the mechanical equilibrium equations to simulate excitation–contraction coupling and its effects on ventricular deformation and pressure–volume behaviour.

### 2.2. Modelling stage-specific HCM phenotypes

Disease-specific alterations were introduced into the healthy-control model using a staged strategy to separate primary sarcomeric dysfunction from secondary ionic, anatomical, and mechanical remodelling.

HCM pathophysiology was initiated by expressing across the entire myocardium the pathogenic MYH7^R403Q/+^ variant, which affects β-myosin heavy chain and is associated with hypercontractility and impaired relaxation. Consistent with prior experimental evidence, the mutation was represented as destabilisation of the myosin super-relaxed state, increasing myosin head availability for cross-bridge formation^8^. This was implemented in the myofilament component of the ToR-ORd–Land framework by modulating cross-bridge recruitment and detachment kinetics, producing enhanced and prolonged active tension without direct modification of baseline ionic current function^36^. The resulting cellular-scale effects of the MYH7 mutation, with and without secondary ionic remodelling, are illustrated in Figure 1A through simulated action potential, calcium transient, and active tension traces.

**Figure 1.**
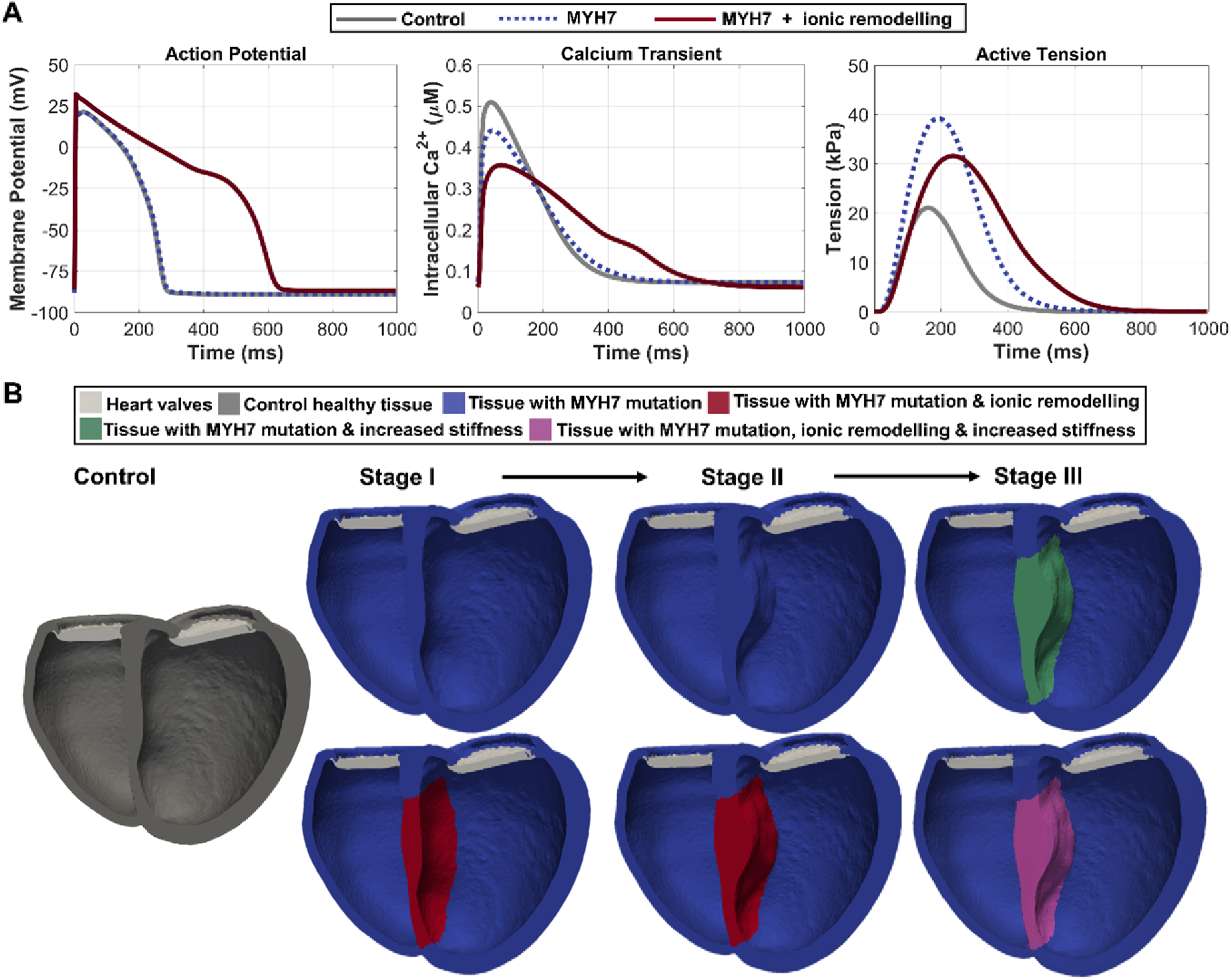
Modelled stages of HCM. (A) Simulated action potential, intracellular calcium transient, and active tension for Control, MYH7 mutation alone, and MYH7 with full (extreme) ionic remodelling. (B) Biventricular geometries illustrating the staged remodelling framework: Stage I, early pre-hypertrophic MYH7 dysfunction; Stage II, septal hypertrophy; and Stage III, septal hypertrophy with increased myocardial stiffness. Each stage was simulated with and without regional ionic remodelling. Colours indicate modelled tissue regions: control myocardium, MYH7-mutant tissue, ionic-remodelled tissue, stiffened myocardium, combined ionic/stiffness remodelling, and heart valves.

Localised secondary ionic remodelling was introduced to account for HCM-associated abnormalities in cellular electrophysiology and calcium handling, based on experimental characterisation of human cardiomyocytes from septal myectomy samples of patients with advanced disease^9,10^. The complete remodelling phenotype was used to represent maximal (extreme) severity^9,37^. Additional moderate and severe ionic-remodelling profiles were simulated to assess severity-dependent effects (see later Figure 5), following previously described definitions^38^; the corresponding conductance changes are detailed in Supplementary Table S1 of that study. Moderate remodelling comprised upregulation of late sodium current alone, whereas severe remodelling included all ionic-remodelling components, with potassium-current changes applied at half the extreme severity^38^.

Ventricular-scale structural remodelling was introduced in three stages, starting from the healthy biventricular geometry (Figure 1B). Stage I represented pre-hypertrophic disease, solely driven by MYH7 dysfunction. Stage II incorporated asymmetric septal hypertrophy as the most common morphological expression of sarcomeric HCM, introduced by geometrically modifying the healthy biventricular mesh to increase maximum interventricular septal thickness from 9.4 mm to approximately 15 mm, representing an approximately 59.6% increase (5.6 mm). Stage III represented advanced adverse remodelling, combining septal hypertrophy with a threefold increase in passive myocardial stiffness across the isotropic, fibre, sheet, and fibre–sheet components of the Holzapfel–Ogden formulation, to represent the increased myocardial stiffness associated with later-stage diastolic dysfunction^39^.

To account for uncertainty in the temporal relationship between electrophysiological and structural remodelling in HCM progression, each structural stage was also simulated with regional septal ionic remodelling. Co-localisation of electrical and structural abnormalities was motivated by previous studies^17,38,40–42^ demonstrating their joint contribution to characteristic ECG manifestations of HCM.

### 2.3. Calibration, validation, and credibility assessment

The model was calibrated under healthy-control conditions and subsequently assessed against clinically measurable biomarkers of ventricular electrophysiology, haemodynamics, and myocardial deformation. The credibility assessment was guided by established verification, validation, and uncertainty-quantification principles for biomedical computational models, with emphasis on the intended context of use and relevant quantities of interest^24,43^. The intended context of use was mechanistic investigation of stage-specific phenotypes in MYH7-related non-obstructive HCM.

The primary quantities of interest included QT interval and QT dispersion (QTd) derived from simulated 12-lead pseudo-ECGs; pressure–volume biomarkers including end-diastolic volume (EDV), end-systolic volume (ESV), stroke volume (SV), EF, and peak systolic pressure (SP); and deformation biomarkers including global longitudinal strain (GLS), global radial strain (GRS), and global circumferential strain (GCS), together with regional septal strain-curve features. End-diastolic pressure (EDP) was prescribed as a stage-specific haemodynamic boundary condition rather than treated as an independently predicted model output. These quantities of interest were compared with literature-derived control and HCM reference ranges^11,12,44,45^, summarised in Table ***1***. Myocardial deformation was additionally evaluated against *in vivo* DENSE-derived strain measurements in healthy subjects as an independent comparator for strain timing and deformation patterns^46^.

**Table 1.** Electrophysiological, haemodynamic, and deformation biomarkers across staged HCM progression. Literature values are shown for control and HCM; model results are shown for control and Stages I–III, representing MYH7 sarcomeric dysfunction, additional septal hypertrophy, and additional myocardial stiffness, respectively. Values are reported as mean ± standard deviation where indicated; model-derived haemodynamic results are single-simulation values. Model-derived QRS duration (QRSd) and corrected QT interval (QTc) are averaged across ECG leads. QT dispersion (QTd) represents the difference between the maximum and minimum uncorrected QT intervals across ECG leads. Model EDP values are prescribed stage-specific loading conditions, not independently predicted outcomes. Abbreviations: QTd, QT dispersion; EDV, end-diastolic volume; ESV, end-systolic volume; SV, stroke volume; EF, ejection fraction; EDP, end-diastolic pressure; SP, peak systolic pressure; GLS, global longitudinal strain; GRS, global radial strain; GCS, global circumferential strain. Model-derived HCM results correspond to the extreme secondary ionic-remodelling profile.

| Biomarker | Literature Values |  | Model Results |  |  |  |
|---|---|---|---|---|---|---|
|  |  |  |  | HCM |  |  |
|  | Control | HCM | Control | Stage I | Stage II | Stage III |
| **Electrophysiological Biomarkers** |  |  |  |  |  |  |
| QRSd (ms) | 94 ± 8<sup>44</sup> | 98±13<sup>44</sup> | 89.4±9.7 | 98.6±12.5 | 109.7±14.8 | 112.5±15.9 |
| QTc (ms) | 413±27<sup>44</sup> | 451±50<sup>44</sup> | 354.7±6.0 | 519.5±22.4 | 541.0±18.5 | 547.9±22.8 |
| QTd (ms) | 46 ± 16<sup>47</sup> | 62 ± 31<sup>47</sup> | 40.3 ± 5.5 | 73.4 ± 0.1 | 78.4 ± 1.2 | 78.8 ± 0.5 |
| **Haemodynamic Biomarkers** |  |  |  |  |  |  |
| EDV (mL) | 140±36<sup>11</sup> | 123±28<sup>11</sup> | 142.7 | 124.8 | 121.3 | 113.9 |
| ESV (mL) | 57±16<sup>11</sup> | 50±18<sup>11</sup> | 56.5 | 45.7 | 41.5 | 42.3 |
| SV (mL) | 83±22<sup>11</sup> | 73±19<sup>11</sup> | 86.2 | 79.1 | 79.8 | 71.6 |
| EF (%) | 59 ± 4<sup>11</sup> | 59±7<sup>11</sup> | 60.4 | 63.4 | 65.8 | 62.9 |
| EDP (mmHg) | 9±3<sup>45</sup> | 15±5<sup>45</sup> | 9.0 | 10.5 | 13.5 | 18.0 |
| SP (mmHg) | 117±11<sup>45</sup> | 131±32<sup>45</sup> | 106.3 | 108.2 | 113.7 | 116.9 |
| **Deformation Biomarkers** |  |  |  |  |  |  |
| GLS (E<sub>ll</sub>, %) | −20.5±2.6<sup>12</sup> | −16.9±2.4<sup>12</sup> | -17.9±0.5 | -15.6±1.2 | -14.2±0.1 | -12.4±0.1 |
| GRS (E<sub>rr</sub>, %) | 47.9±7.4<sup>12</sup> | 29 ±7.2<sup>12</sup> | 54.8±0.6 | 39.2±1.1 | 39.6±1.0 | 38.2±1.1 |
| GCS (E<sub>cc</sub>, %) | −20.8±7.4<sup>12</sup> | −17.3±2.6<sup>12</sup> | -18.8±0.1 | -17.3±0.6 | -16.6±0.2 | -17.4±0.4 |

Healthy-control calibration was performed before disease-specific remodelling was introduced. Electrophysiological calibration targeted physiological activation patterns and ECG morphology by adjusting earliest activation sites and conduction properties within the fast endocardial activation layer. Mechanical calibration targeted physiological LV pressure– volume behaviour within established healthy reference ranges by selecting passive material properties, pressure boundary conditions, Windkessel parameters, and pericardial constraint values. Strain metrics were not directly tuned, but were retained as independent deformation-based comparators. End-diastolic pressure and arterial resistance were adjusted across stages to represent haemodynamic changes associated with HCM progression (Supplementary Section 1.5 and Supplementary Table S1). After calibration, staged HCM simulations were evaluated for consistency with independent clinical and imaging biomarkers reported for HCM.

Model credibility was therefore based on the use of established electromechanical solvers and validated cellular and myofilament formulations; calibration of the healthy-control model before disease remodelling; comparison of staged HCM outputs with independent clinical and imaging biomarkers; and the staged perturbation design, which allowed sarcomeric dysfunction, ionic remodelling, hypertrophy, and increased stiffness to be examined individually and in combination.

## 3. Results

### 3.1. Healthy-control model reproduced physiological electromechanical function

The calibrated healthy-control model reproduced physiological electrical activation, ventricular haemodynamics, ECG morphology, and myocardial deformation (Figure 2 and Figure 3, Supplementary Video S1). Ventricular activation followed a physiological septum-to-apex-to-base sequence (Figure 2A), generating physiological QRS durations and normal T-wave morphology in the simulated precordial ECG leads (Figure 2B).

**Figure 2.**
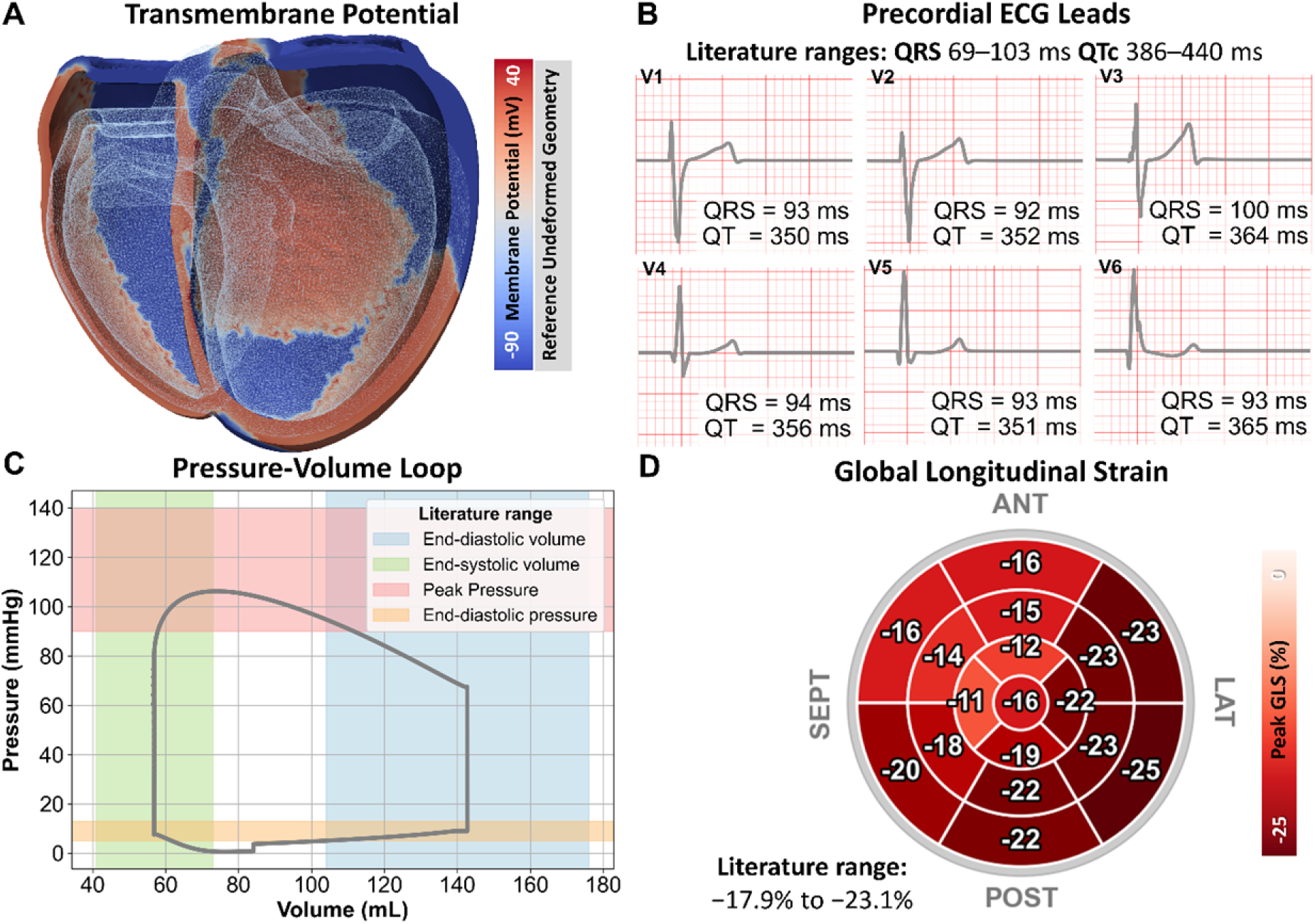
Multiscale electrophysiological, haemodynamic, and deformation biomarkers in the calibrated healthy-control. (A) Spatial distribution of transmembrane potential during QRS-phase ventricular activation, shown on the biventricular geometry, with the undeformed reference geometry overlaid in grey. (B) Simulated precordial ECG leads V1–V6. (C) Left ventricular pressure–volume loop, with shaded regions indicating physiological reference ranges. (D) Peak systolic global longitudinal strain (GLS) displayed using the 17-segment American Heart Association bullseye model. Segmental strain values are shown as percentages.

**Figure 3.**
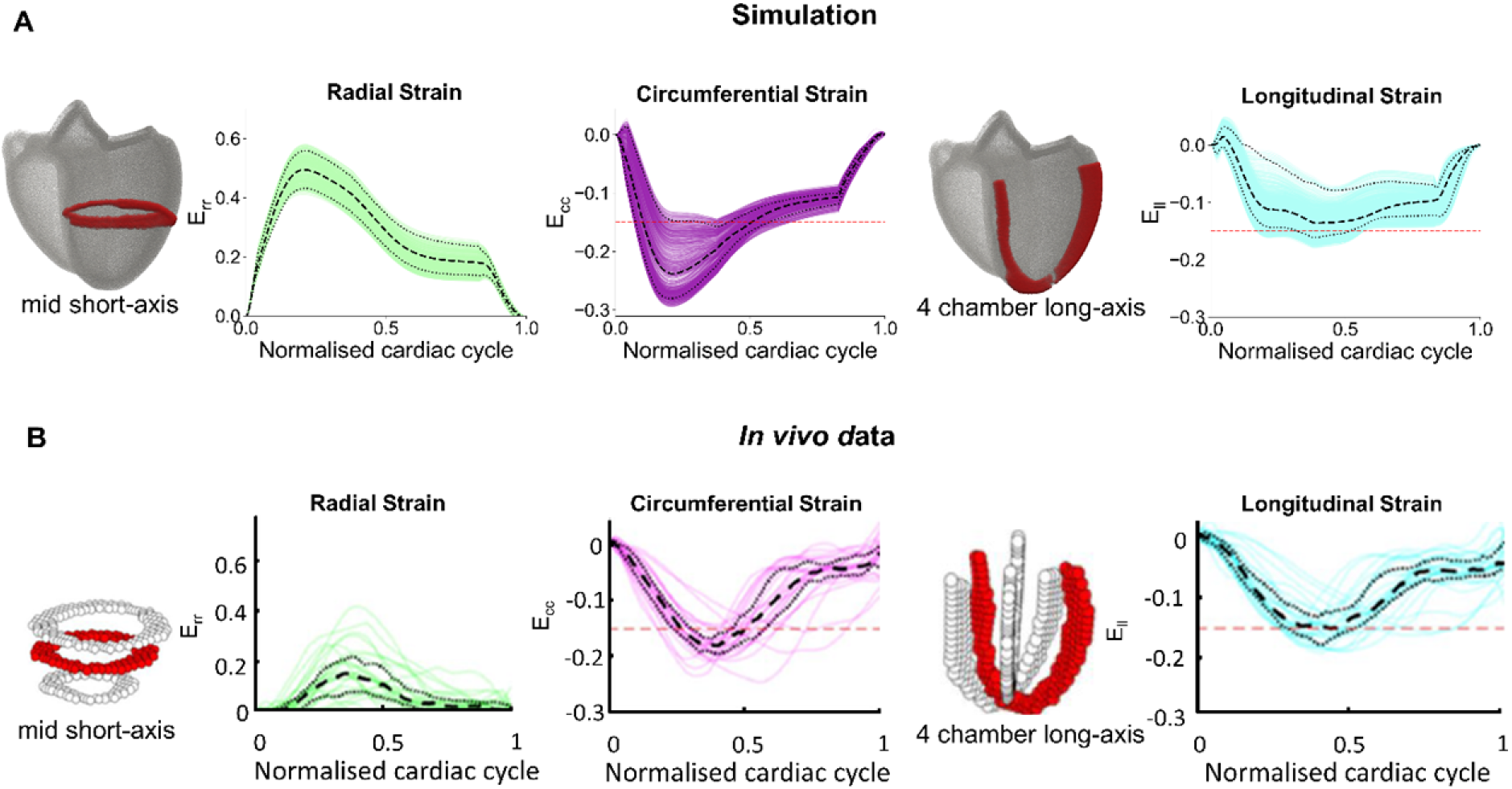
Comparison of simulated and *in vivo* left ventricular strain in healthy hearts. (A) Simulated radial (Err), circumferential (Ecc), and longitudinal (Ell) strain, normalised to the cardiac cycle duration. Strain was sampled from the highlighted short-axis and long-axis regions. Dashed lines indicate median values and dotted lines the interquartile range, reflecting spatial heterogeneity within the simulated ventricles. (B) Corresponding *in vivo* strain data obtained using DENSE in 30 healthy subjects. Thin lines represent individual subjects, with dashed lines indicating the cohort median and dotted lines the interquartile range, reflecting inter-subject variability. The horizontal dashed reference lines denote a strain value of −0.15. Panel B was adapted from Figure 6 in Moulin et al.^46^ with permission from Wiley.

At the chamber level, the model exhibited physiological pressure–volume behaviour during filling, ejection, and relaxation (Figure 2C). EDV was 142.7 mL, ESV was 56.5 mL, and EF was 60.4%. Peak LV pressure reached 106.3 mmHg, and EDP was prescribed at 9 mmHg. Regional strain showed spatial heterogeneity, with greater shortening in the lateral and posterior walls and lower strain in the septal and apical regions (Figure 2D).

Myocardial deformation was further evaluated against *in vivo* DENSE-derived strain measurements reported by Moulin et al.^46^ in 30 healthy subjects (Figure 3). For this comparison, simulated strain was sampled from the slice nodes highlighted in Figure 3A: the mid short-axis slice for radial and circumferential strain, and the four-chamber long-axis slice for longitudinal strain. These slice-based values are therefore distinct from the global deformation biomarkers reported in

Table ***1***. Strain values are reported as dimensionless median [interquartile range] at peak systolic deformation. Simulated radial strain increased during systole to approximately 0.49 [0.43, 0.55], circumferential strain reached approximately −0.23 [−0.28, −0.16], and longitudinal strain reached approximately −0.13 [−0.16, −0.1]. The *in vivo* cohort showed median radial, circumferential, and longitudinal strain values of approximately 0.12–0.16, −0.17 to −0.19, and −0.13 to −0.15, respectively. Circumferential and longitudinal strain were therefore broadly consistent with the corresponding i*n vivo* ranges, whereas simulated radial strain was substantially greater than DENSE-measured radial strain. However, Moulin et al. reported that DENSE underestimates radial strain relative to ground truth in computational phantom validation^46^, which should be considered when interpreting this discrepancy.

### 3.2. Sarcomeric dysfunction initiated a low-volume hypercontractile phenotype

Relative to the healthy control, Stage I without ionic remodelling reduced EDV from 142.7 to 136.1 mL and ESV from 56.5 to 48.5 mL, while increasing EF from 60.4% to 64.3% and peak LV pressure from 106.3 to 113.8 mmHg (Figure 4A). The Stage I phenotype was therefore characterised by reduced ventricular volumes and increased EF before the development of overt hypertrophy, while stroke volume remained preserved.

**Figure 4.**
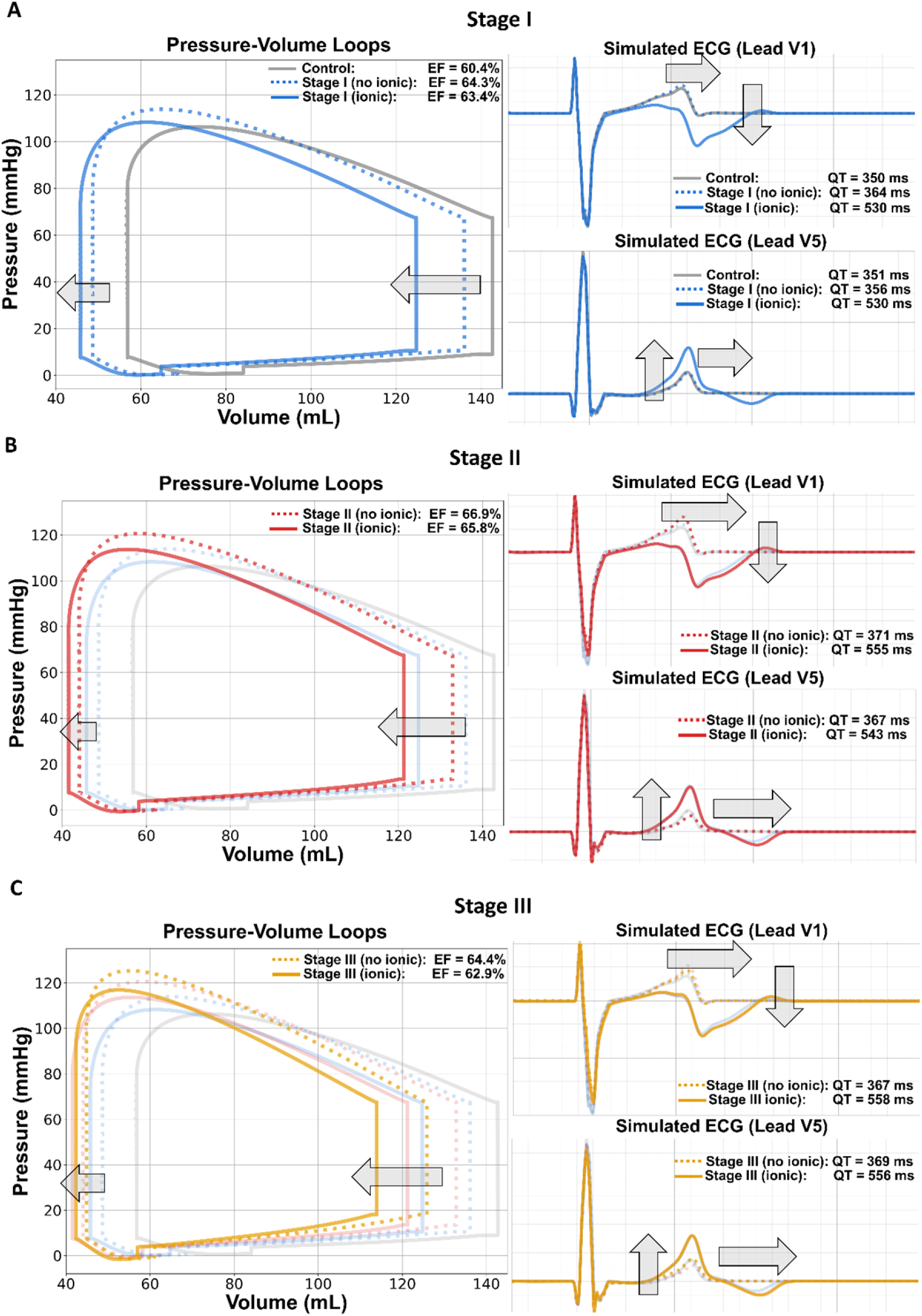
Pressure–volume and ECG changes across HCM stages. Simulated left ventricular pressure–volume loops and ECG traces (leads V1 and V5) are shown for the healthy control case and progressive HCM stages: (A) MYH7-driven sarcomeric dysfunction (Stage I), (B) septal hypertrophy (Stage II), and (C) septal hypertrophy with increased myocardial stiffness (Stage III). Dotted and solid traces denote simulations without ionic remodelling and with the extreme secondary ionic-remodelling profile, respectively. Ejection fraction (EF) and QT interval are annotated for each condition. Grey and faded coloured traces indicate the control and preceding stages for visual comparison. Across the staged HCM phenotypes, pressure–volume loops shift towards lower ventricular volumes. Secondary ionic remodelling further reduces ventricular filling and is accompanied by marked QT prolongation and T-wave abnormalities.

**Figure 5.**
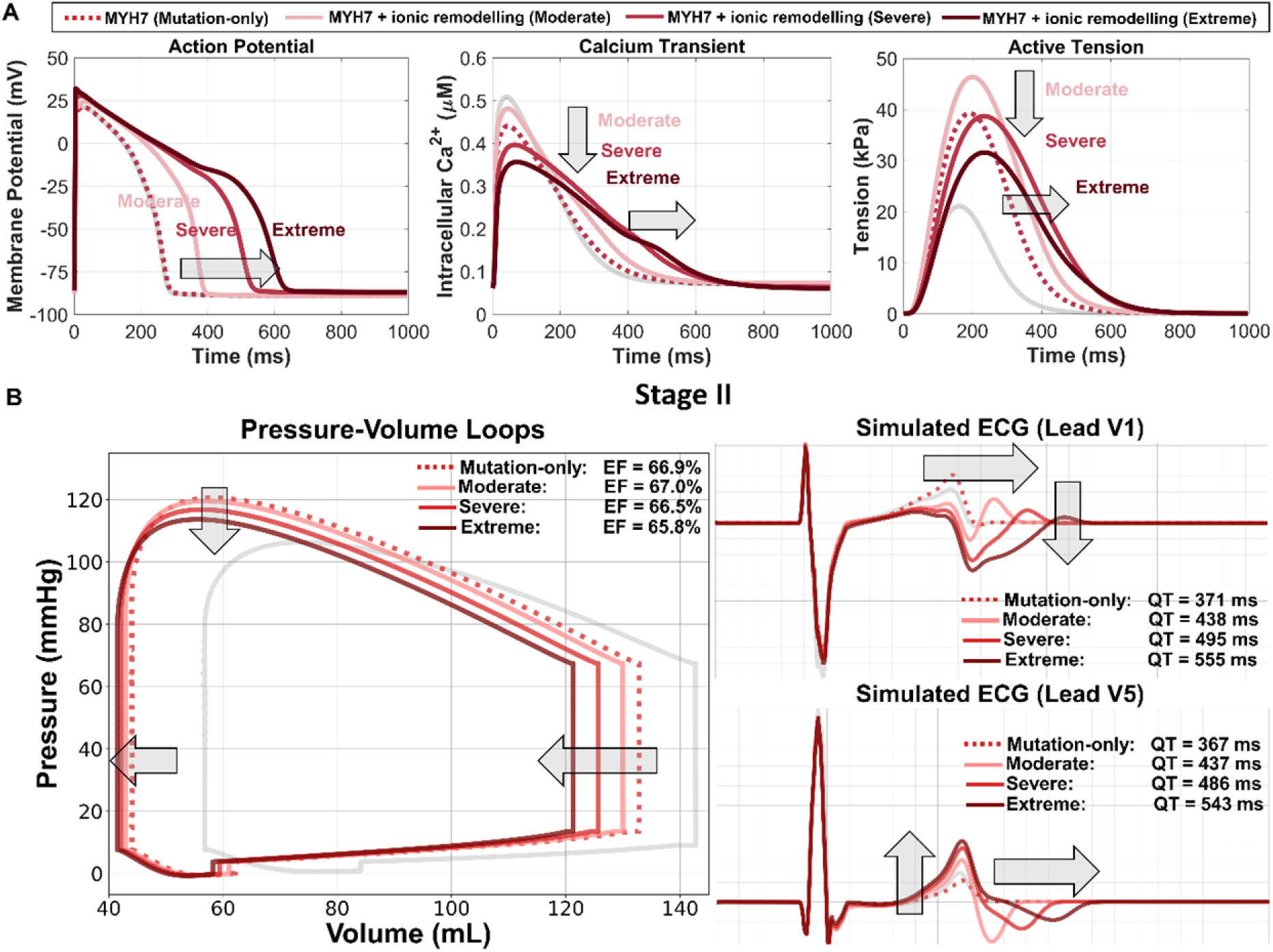
Sensitivity of pressure-volume loops and ECGs to graded secondary ionic remodelling. (A) Simulated cellular action potential, intracellular calcium transient, and active tension for MYH7 mutation alone and increasing ionic *remodelling severity. (B) Corresponding* pressure–volume loops and ECG traces (leads V1 and V5) in Stage II HCM. Ejection fraction and QT interval are annotated.

### 3.3. Structural remodelling progressively constrained ventricular filling

In Stage II without ionic remodelling, EDV decreased from 136.1 to 132.9 mL and ESV from 48.5 to 44.0 mL relative to Stage I, while EF increased to 66.9% and peak LV pressure to 120.5 mmHg (Figure 4B). Stroke volume remained similar to Stage I (88.9 versus 87.6 mL) despite the lower EDV. Thus, the Stage II phenotype retained the low-volume hypercontractile phenotype initiated by sarcomeric dysfunction, with higher EF and peak LV pressure accompanying preservation of stroke volume despite further chamber constraint.

Stage III, incorporating increased myocardial stiffness without ionic remodelling (Figure 4C), showed a further reduction in ventricular volumes, with EDV decreasing to 125.9 mL, while peak LV pressure increased to 125.3 mmHg and EF remained elevated at 64.4%. Stroke volume decreased to approximately 81.1 mL, indicating that the Stage III phenotype combined reduced filling and stroke volume with preserved EF.

Across stages without ionic remodelling, ventricular volumes decreased as sarcomeric dysfunction, septal hypertrophy, and increased myocardial stiffness were introduced sequentially. Although EF remained elevated throughout, stroke volume was maintained near control levels through Stage II before declining in Stage III. These findings demonstrate that preserved EF can coexist with reduced ventricular filling and stroke volume during advanced HCM remodelling.

### 3.4. Secondary ionic remodelling prolonged repolarisation and exacerbated diastolic dysfunction

Secondary ionic remodelling produced the most pronounced repolarisation effects in the staged disease framework (Figure 4). In the absence of ionic remodelling, QT interval changed only modestly across sarcomeric and structural disease progression. By contrast, secondary ionic remodelling resulted in clinically relevant QT prolongation (>480 ms) at all stages, accompanied by T-wave abnormalities, consistent with the expected HCM repolarisation phenotype. QT dispersion also increased from 40.3±5.5 ms in control to 73.4±0.1, 78.4±1.2, and 78.8±0.5 ms in Stages I–III with extreme ionic remodelling, respectively (Table ***1***), indicating greater inter-lead variation in QT duration. QRS changes were comparatively small and were therefore not interpreted as a primary mechanistic effect. These findings identify delayed repolarisation and increased QT dispersion as the dominant electrical consequences of secondary ionic remodelling.

Beyond this established electrophysiological phenotype, the main organ-scale effect of ionic remodelling was impaired ventricular filling. Within each disease stage, ionic-remodelling effects were assessed using the same ventricular geometry, passive myocardial properties, and haemodynamic boundary conditions. Ionic remodelling further reduced ventricular volumes and shifted pressure–volume loops leftwards at each disease stage, while preserving EF. Across stages, ionic remodelling reduced EDV by 11.6 mL on average. In Stage I, ionic remodelling reduced EDV from 136.1 to 124.8 mL and ESV from 48.5 to 45.7 mL, while EF remained elevated at 63.4%. In Stage II, EDV decreased from 132.9 to 121.3 mL and ESV from 44.0 to 41.5 mL, with EF remaining 65.8%. In Stage III, the combination of stiffness and ionic remodelling produced the smallest ventricular volumes, with EDV of 113.9 mL and ESV of 42.3 mL, while EF remained preserved at 62.9%. Despite the preserved EF, stroke volume decreased to 71.6 mL, compared with approximately 81.1 mL in Stage III without ionic remodelling, indicating a reduction in forward volume that was not apparent from EF alone.

Notably, secondary ionic remodelling consistently reduced ventricular filling across all three disease stages (Figure 4; Supplementary Figure S2). The accompanying prolongation of calcium–tension dynamics and delayed relaxation (Figure 5A) provide a mechanistic explanation for these reductions in ventricular volume.

### 3.5. Graded ionic remodelling produced severity-dependent electrical and mechanical abnormalities

To further isolate the contribution of secondary ionic remodelling, moderate, severe, and extreme remodelling profiles were examined across cellular and organ scales (Figure 5). This analysis tested whether the ionic-remodelling effects described above were preserved across different remodelling severities.

At the cellular level (Figure 5A), relative to MYH7 mutation-only, increasing ionic remodelling severity produced progressive prolongation of action potential and calcium transient duration. In contrast, peak intracellular calcium exhibited a biphasic response, increasing first with moderate ionic remodelling, consistent with enhanced late sodium current and secondary calcium loading, then decreasing at higher severities owing to progressive intracellular calcium handling remodelling. Peak active tension showed a similar biphasic response. Despite lower peak force at higher severities, contraction duration increased from 568 ms with MYH7 mutation alone to 620, 696, and 708 ms with moderate, severe, and extreme remodelling, respectively. The tension–time integral remained above the mutation-only value of 9.51 kPa·s, reaching 12.62, 12.56, and 10.30 kPa·s with moderate, severe, and extreme remodelling, respectively.

At organ scale, graded ionic remodelling progressively reduced ventricular volumes and modestly attenuated peak systolic pressure while preserving EF (see Figure 5B for the representative Stage II hypertrophic phenotype; Supplementary Figure S2 for consistent findings across the remaining structural-remodelling stages). The ECG response was dominated by severity-dependent QT prolongation, whereas QRS changes were small and lead-dependent. QT dispersion also generally increased with ionic-remodelling severity across the three stages (Supplementary Table S2), suggesting greater variability in repolarisation timing between ECG leads.

Across scales, increasing ionic-remodelling severity therefore predominantly prolonged repolarisation and calcium–tension dynamics. These changes occurred without proportional increases in peak contractile force or consistent conduction delay, indicating impaired relaxation rather than enhanced systolic force generation as the principal mechanical consequence.

### 3.6. Integrated biomarkers revealed progressive disease burden despite preserved ejection fraction

Integrated biomarker analysis showed progressive electromechanical deterioration across HCM stages despite preserved EF (Table ***1***). The main results in Table ***1*** correspond to the extreme secondary ionic-remodelling profile, whereas Supplementary Table S2 reports results across all ionic-remodelling severities, including mutation-only, moderate, severe, and extreme remodelling, for each disease stage. Comparison with literature-reported control and HCM ranges showed broad consistency of the staged simulations with reported electrical, haemodynamic, and deformation phenotypes. Electrical remodelling was characterised by QTc prolongation, consistent with delayed repolarisation. Haemodynamic indices showed progressively reduced ventricular volumes and lower stroke volume in advanced disease. Although EF remained within the normal range throughout disease progression, global strain metrics revealed worsening myocardial deformation not captured by EF alone, most prominently through impaired longitudinal shortening and reduced radial strain, with circumferential strain also becoming abnormal relative to control.

Together, these findings show that preserved EF did not preclude progressive electrical, haemodynamic, and mechanical abnormalities. Repolarisation, ventricular volumes, and myocardial deformation provided complementary indicators of electromechanical disease burden.

### 3.7. Septal strain and active-stress maps identified regional mechanical dysfunction

Regional septal strain analysis provided regional readouts of stage-dependent mechanical deterioration (see Figure 6A for full ionic remodelling, with other remodelling severities presented in Supplementary Figure S3). Circumferential strain showed progressive attenuation across HCM stages, with peak strain becoming progressively less negative, from −18.7% in Stage I to −15.0% in Stage II and −12.1% in Stage III, indicating reduced circumferential shortening. Late-diastolic slope declined from 75.7 to 63.2 and 38.7, respectively, indicating progressive blunting of late-diastolic recovery. Radial strain showed a marked reduction relative to control, with peak radial strain falling from 72.2% in control to 38.9%, 31.8%, and 24.2% across Stages I–III. The corresponding late-diastolic slope decreased from 350.9 in control to 107.0, 71.8, and 63.7, respectively. Longitudinal strain was similarly impaired across all disease stages relative to control, with peak shortening attenuated from −20.2% in control to −12.6%, −12.8%, and −9.7% in Stages I–III, respectively, and late-diastolic slope declining from 114.7 to 76.0, 74.8, and 58.3. Across all strain components, the staged HCM phenotypes showed attenuated septal shortening or radial deformation and impaired late-diastolic recovery.

**Figure 6.**
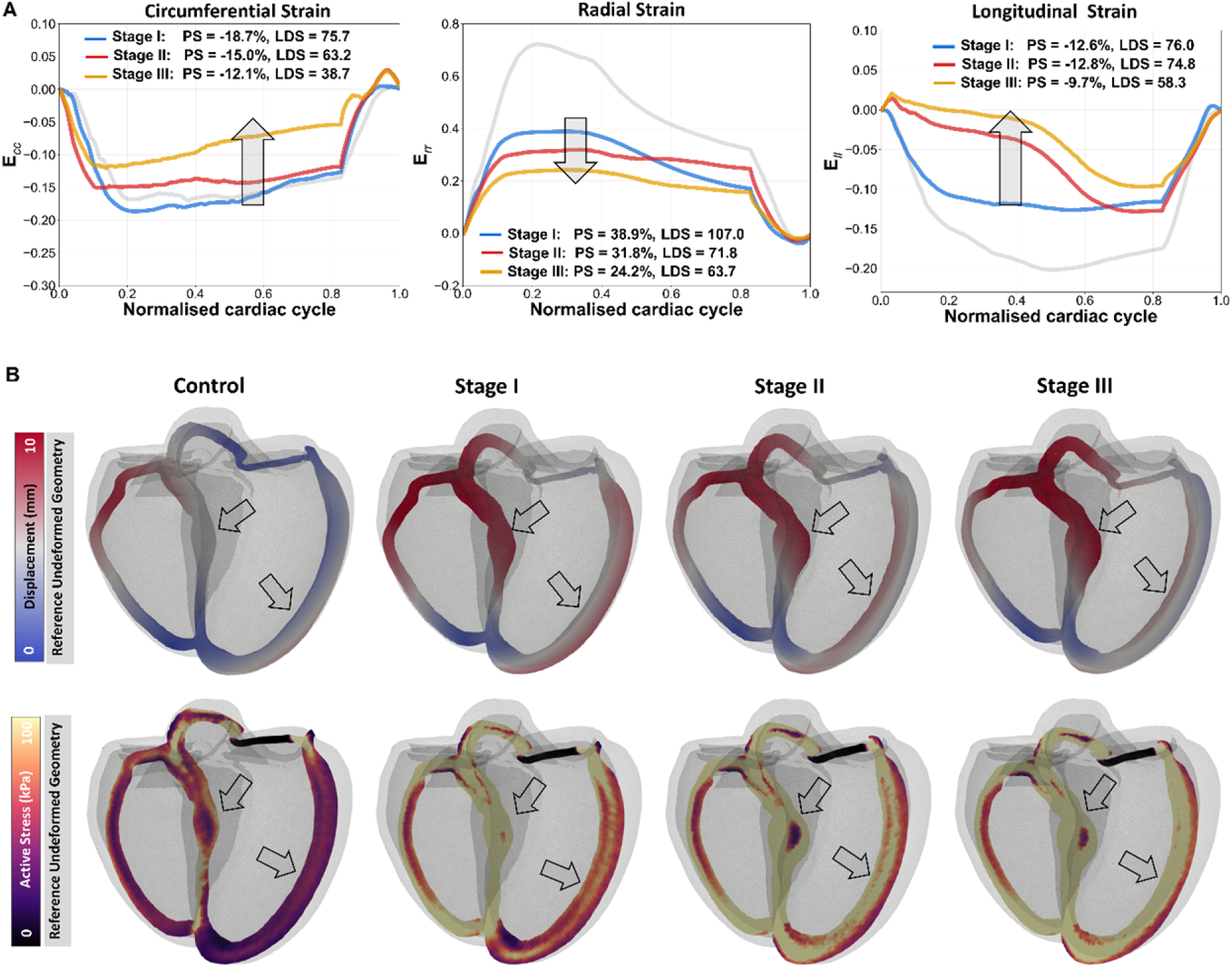
Regional strain abnormalities and mechanical heterogeneity across HCM progression. (A) Median septal circumferential (E_cc_), radial (E_rr_), and longitudinal (E_ll_) strain throughout the normalised cardiac cycle for Stage I, Stage II, and Stage III HCM under the extreme ionic-remodelling profile; the grey trace denotes the healthy-control reference. Peak strain (PS) and late-diastolic slope (LDS) are annotated for each stage. Disease progression is associated with attenuated septal shortening or thickening and blunted late-diastolic recoil. (B) End-systolic displacement magnitude (top) and active-stress distributions (bottom) in the healthy-control and staged HCM phenotypes. Reference undeformed geometries are shown in grey for anatomical reference. Arrows indicate corresponding anatomical regions across stages, highlighting progressive changes in the same septal and adjacent myocardial regions. Progressive disease produces increasing regional mechanical heterogeneity and focal active-stress localisation within the septum and adjacent myocardium.

Regional displacement and active-stress distributions revealed increasing mechanical heterogeneity across HCM stages (Figure 6B). Whereas the healthy-control model exhibited relatively smooth end-systolic displacement patterns, the HCM stages showed increasingly heterogeneous displacement, with regional accentuation in the septal and adjacent ventricular wall regions (Supplementary Video S2). These changes reflected redistribution of regional motion within a preserved global systolic phenotype rather than changes that could be inferred from EF alone. Active-stress showed a distinct stage-dependent pattern, transitioning from a relatively uniform distribution across the ventricular wall in the healthy-control model to focal elevations of stress in the septum and adjacent myocardium across HCM stages. The accompanying changes in the free wall likely reflect biventricular mechanical coupling and redistribution of load, rather than an isolated septal response. These abnormalities became most pronounced in Stage III, consistent with greater regional mechanical demand after the addition of myocardial stiffening despite preserved EF. Spatiotemporal analysis of transmembrane potential and displacement showed delayed repolarisation during ejection in the presence of ionic remodelling, accompanied by greater temporal separation between electrical recovery and mechanical deformation in the hypertrophic and stiffened stages (Supplementary Figure S4).

Together, these results show that regional strain and active-stress distributions captured stage-dependent mechanical abnormalities that were not apparent from EF alone. In particular, the staged HCM phenotypes developed increasing regional mechanical heterogeneity despite preserved global systolic function.

## 4. Discussion

In this study, we developed a human biventricular electromechanical framework to investigate the mechanisms contributing to diastolic dysfunction across staged phenotypes of non-obstructive HCM, linking genotype-related cellular dysfunction to organ-scale phenotype. The principal mechanistic insight is that no single disease component accounted for the full HCM phenotype; rather, distinct sarcomeric, electrophysiological, structural, and mechanical remodelling processes contributed complementary effects. Sarcomeric dysfunction initiated an early low-volume, hypercontractile phenotype; secondary ionic remodelling prolonged repolarisation and calcium–tension dynamics, thereby impairing relaxation; septal hypertrophy imposed chamber-level mechanical constraint and altered deformation; and the Stage III phenotype, incorporating increased myocardial stiffness, showed further reductions in filling and greater regional mechanical heterogeneity. Importantly, these changes occurred despite preserved EF, indicating that EF alone may underestimate the extent of electromechanical disease burden. Together, the findings highlight the complementary roles of sarcomeric dysfunction, secondary ionic remodelling, septal hypertrophy, and increased myocardial stiffness in shaping the evolving electromechanical phenotype of HCM.

### 4.1. Mechanistic interpretation of staged HCM progression

Diastolic dysfunction in HCM is multifactorial, involving mechanisms that extend beyond hypertrophy alone. The staged modelling framework links MYH7-mediated sarcomeric dysfunction to the organ-scale HCM phenotype while examining the contributions of septal hypertrophy, increased myocardial stiffness, and secondary ionic remodelling to ventricular filling and myocardial deformation. At the earliest stage, destabilisation of the myosin super-relaxed state was associated with a low-volume, hypercontractile phenotype characterised by reduced ventricular volumes, increased EF, and elevated peak systolic pressure. This is consistent with experimental evidence that abnormal contractile regulation is an early feature of HCM and can precede overt systolic pump failure^8^. Importantly, the Stage I phenotype developed before the introduction of septal hypertrophy or secondary ionic remodelling, highlighting the early involvement of sarcomeric dysfunction in HCM phenotype expression.

Secondary ionic remodelling produced a distinct mechanistic signature. The predominant effects identified here were prolonged repolarisation and calcium–tension dynamics together with impaired ventricular filling. Increasing ionic-remodelling severity produced progressive prolongation of repolarisation and calcium-tension dynamics, while reducing ventricular volumes without proportionally increasing peak calcium or peak contractile force. This pattern supports impaired relaxation, rather than augmented peak systolic force generation, as the principal mechanical consequence of secondary ionic remodelling. Notably, ionic remodelling reduced EDV by approximately 11–12 mL within each disease stage, a magnitude comparable to the 10.2 mL reduction observed between Stages I and III without ionic remodelling. Together with the associated prolongation of calcium–tension dynamics, these findings support a substantial contribution of secondary ionic abnormalities to impaired ventricular relaxation and filling, beyond their effects on repolarisation.

Structural remodelling was associated with further reductions in ventricular filling within the low-volume phenotype established at Stage I. The Stage II phenotype showed lower EDV with preserved stroke volume, whereas Stage III, incorporating increased myocardial stiffness, exhibited a further reduction in EDV accompanied by lower stroke volume. These changes were also associated with greater regional mechanical heterogeneity. Together, these findings are consistent with the clinical observation that HCM can exhibit worsening diastolic and regional myocardial dysfunction despite preserved EF^1–4^. In this setting, preserved EF reflects fractional emptying of a progressively smaller ventricle and should not be interpreted as evidence of preserved stroke volume or myocardial function. The relative contributions of these mechanisms are likely to vary with hypertrophy pattern, genetic background, treatment, and comorbid disease, motivating further evaluation across broader and patient-specific HCM phenotypes.

### 4.2. Regional biomarkers reveal disease burden beyond ejection fraction

Regional strain and active-stress analyses provided mechanistic readouts of myocardial dysfunction beyond chamber-level haemodynamic indices. Global deformation biomarkers and septal strain features showed stage-dependent mechanical impairment, indicating worsening myocardial function. These findings are consistent with clinical studies showing that strain imaging can reveal subclinical or regional dysfunction despite preserved EF^11,12^. In particular, septal strain indices, including peak strain and late-diastolic slope, captured stage-dependent mechanical deterioration and delayed recovery (Figure 6). Graded ionic remodelling further amplified mechanical dysfunction across the cardiac cycle, particularly through reduced radial strain, impaired longitudinal shortening, and blunted late-diastolic strain recovery (Supplementary Figure S3).

Active-stress mapping provided additional information on regional force generation that is not directly measurable *in vivo*. Unlike strain, pressure, or displacement, which can be assessed clinically, active myocardial stress is a model-derived quantity that enables spatial and temporal localisation of mechanical demand within the ventricular wall. Across the staged HCM phenotypes, active-stress distributions became increasingly heterogeneous, with focal elevations emerging in the septum and adjacent myocardium. These changes were accompanied by greater electromechanical heterogeneity, with delayed repolarisation and impaired relaxation contributing to regional dysfunction (Supplementary Figure S4).

Together, these findings support regional strain as a clinically accessible marker of mechanical dysfunction, while active-stress mapping provides a complementary mechanistic readout of regional myocardial force generation. Both revealed abnormalities in deformation, recovery, and mechanical demand that were not apparent from EF alone.

### 4.3. Clinical and translational implications

Our findings reinforce that preserved EF is an incomplete marker of disease burden in non-obstructive HCM. Despite preserved EF, the staged HCM phenotypes showed worsening ventricular filling, repolarisation abnormalities, impaired myocardial deformation, and increasing regional mechanical heterogeneity. These observations support the use of complementary clinical readouts, including ECG, pressure–volume, and strain-based measures, alongside model-derived active-stress information, to provide a more comprehensive assessment of electromechanical disease burden.

More broadly, the staged framework associated distinct readouts with different components of the HCM phenotype. Pressure–volume indices captured low-volume hypercontractility and chamber constraint, QT prolongation reflected secondary ionic remodelling, and strain captured regional mechanical deterioration, while active-stress mapping provided a complementary model-derived measure of regional mechanical demand. As disease-modifying therapies increasingly target specific pathways within the HCM cascade, mechanistic frameworks such as this may help relate measurable biomarkers to underlying disease mechanisms and inform future approaches to disease stratification and therapeutic evaluation^2,9,48^.

### 4.4. Limitations

This study should be interpreted in light of several limitations. The framework was designed for mechanistic investigation rather than patient-specific prediction and, although the healthy-control model was calibrated and the staged HCM outputs were compared with clinically measurable reference biomarkers, the staged HCM phenotypes represent mechanistically defined disease states rather than longitudinal trajectories within individual patients. Stage-specific end-diastolic pressure and arterial resistance were adjusted to reflect reported haemodynamic changes during HCM progression. Accordingly, between-stage pressure– volume differences reflect both myocardial remodelling and the prescribed haemodynamic environment, whereas within-stage ionic-remodelling comparisons were performed under identical boundary conditions. Myocardial fibre disarray and fibrosis were not explicitly represented as structural substrates. Increased passive stiffness in Stage III captured part of the mechanical phenotype associated with advanced remodelling but did not reproduce these tissue abnormalities explicitly. Atrial mechanics and remodelling were not explicitly represented, and their potential contributions to ventricular filling remain to be investigated. The model was limited to non-obstructive HCM without explicit modelling of dynamic LV outflow tract obstruction or mitral valve–septal interactions. The findings are therefore most directly applicable to MYH7-related, septal-predominant, non-obstructive HCM; generalisability to other hypertrophy phenotypes, medication states, comorbid ischaemia, and other mutations remains to be established. In addition, uncertainty was addressed through staged perturbations rather than formal population-based or sensitivity analysis. Future work should address these limitations through patient-specific calibration, explicit representation of fibre disarray and fibrosis, incorporation of atrial mechanics and remodelling, and rigorous uncertainty quantification.

## 5. Conclusions

Using a staged human biventricular electromechanical model, we show that diastolic dysfunction in HCM arises from distinct but complementary sarcomeric, geometric, stiffness-related, and ionic mechanisms, rather than hypertrophy alone. MYH7 sarcomeric dysfunction initiated a low-volume, hypercontractile phenotype; septal hypertrophy imposed geometric constraint; increased myocardial stiffness further limited ventricular filling; and secondary ionic remodelling prolonged repolarisation and calcium–tension dynamics, thereby impairing relaxation. Despite preserved EF across the staged HCM phenotypes, ventricular filling and regional myocardial mechanics progressively deteriorated, indicating that EF alone does not capture the full electromechanical disease burden. ECG, pressure–volume, and regional strain measures provide complementary clinical readouts, while active-stress mapping offers model-derived mechanistic information on regional myocardial force generation that is not directly accessible from clinical measurements.

## Ethics Statement

This study used previously published and/or de-identified data and did not involve the acquisition of new human participant or animal data. No additional ethical approval was required.

## Data Availability

The simulation data, model configuration files, and analysis scripts underlying this article will be made available by the corresponding author upon reasonable request for scientific investigation.

## CRediT authorship contribution statement

A.I.H.: Conceptualisation, Data curation, Formal analysis, Investigation, Methodology, Software, Validation, Visualisation, Writing – original draft, Writing – review & editing. J.A.C.: Conceptualisation, Methodology, Writing – review & editing. Z.J.W., R.D., X.Z., A.Z., M.V., A.A., R.C., I.O.: Methodology, Writing – review & editing. A.B.-O.: Conceptualisation, Methodology, Funding acquisition, Project administration, Resources, Supervision, Writing – review & editing.

