## Supplementary for "Electromechanical Drivers of Diastolic Dysfunction During Hypertrophic Cardiomyopathy Progression"

##### Supplementary Material

Abdallah I Hasaballa[a], James A Coleman[a], Zhinuo J Wang[a], Ruben Doste[a], Xin Zhou[a], Alberto Zingaro[b], Mariano Vázquez[b,c], Alessia Argirò[d], Raffaele Coppini[e], Iacopo Olivotto[d], Alfonso Bueno-Orovio[a,*]

[a] Department of Computer Science, University of Oxford, Oxford, United Kingdom
[b] ELEM Biotech, Barcelona, Spain
[c] Barcelona Supercomputing Center, Barcelona, Spain
[d] Cardiomyopathy Unit, Careggi University Hospital, Florence, Italy
[e] Department of Preclinical and Clinical Pharmacology, University of Florence, Florence, Italy

### 1. Computational methods and model parameterisation

A human biventricular electromechanical modelling framework was implemented to link cellular disease mechanisms with organ-scale electrical, haemodynamic, and mechanical phenotypes in a calibrated healthy-control model and staged HCM simulations (Supplementary Figure S1). Simulations were performed using Alya, a highly parallel multiphysics simulation code developed at the Barcelona Supercomputing Center[1,2]. The electrical and mechanical subproblems were solved in separate modules and coupled through the electromechanical workflow described in the main Methods. The framework builds on previously established human ventricular electromechanical modelling approaches that integrate cellular electrophysiology, calcium handling, active tension generation, ventricular mechanics, circulatory loading, pressure–volume dynamics, pseudo-ECGs, and myocardial deformation analysis[3–6]. This design enabled systematic evaluation of the MYH7 mutation, septal hypertrophy, increased myocardial stiffness, and secondary ionic remodelling as distinct but interacting contributors to HCM phenotype expression.

#### 1.1. Anatomical model and mesh discretisation

A biventricular finite-element mesh composed of linear tetrahedral elements, with the valvular openings closed to form enclosed ventricular cavities, was derived from four-chamber cardiac computed tomography geometries of a healthy human heart obtained from a published database[7]. The enclosed cavities enabled ventricular pressure loading and chamber-level mechanical simulation. The mesh had a mean equivalent tetrahedral edge length of 1.02 mm. Rule-based fibre, sheetlet, and sheet-normal orientations were assigned throughout the myocardium to define orthotropic electrical conduction and mechanical behaviour[8]. The associated spatial fields and input files required for simulation were generated as part of the same preprocessing workflow.

The control and Stage I simulations used the same biventricular mesh, containing 212,404 nodes, 1,093,149 tetrahedral elements, and 120,328 boundary elements. The Stage II and

Stage III simulations used the hypertrophied biventricular geometry, generated in Blender by locally deforming the interventricular septal surface to reproduce asymmetric septal hypertrophy. The modified surface geometry was subsequently tetrahedralised to generate a volumetric mesh containing 220,698 nodes, 1,143,498 tetrahedral elements, and 120,096 boundary elements. Within each corresponding simulation group, the same mesh was used across conditions to ensure that differences in simulated biomarkers reflected the imposed disease mechanisms.

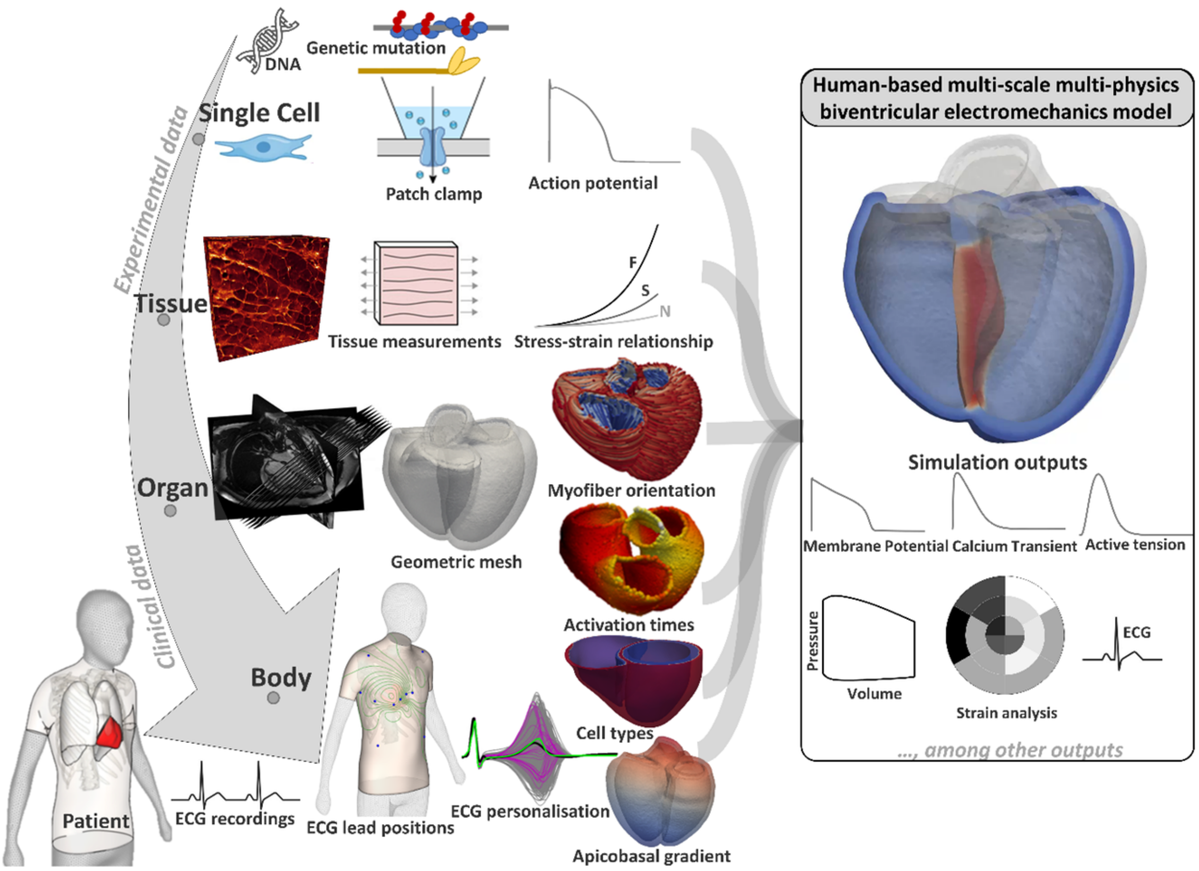


**Supplementary Figure S1. Human-based multiscale biventricular electromechanics framework.** Experimental and clinical data inform cellular electrophysiology, tissue mechanics, and organ-level anatomy. A biventricular finite-element geometry is combined with myocardial microstructure (fibre architecture), activation timing, and physiological heterogeneity (cell types and apico-basal gradients) to run coupled electromechanical simulations. Model outputs include membrane potential, calcium transients, active tension, pressure–volume loops, strain metrics, and pseudo-ECGs, enabling in silico investigation of HCM mechanisms and progression.

#### 1.2. Electrophysiology and ECG computation

Electrical activation and propagation were simulated using the monodomain equation with orthotropic diffusion aligned with the local myocardial fibre, sheetlet, and sheet-normal directions. At the employed mean equivalent tetrahedral edge length of 1.02 mm, diffusion coefficients were selected to reproduce conduction velocities of 67, 30, and 17 cm/s along the fibre, sheetlet, and sheet-normal directions, respectively[9]. Cellular ionic currents and intracellular calcium handling were modelled using the human ToR-ORd ventricular electrophysiological model[10]. Physiological ventricular heterogeneity was represented using

70% endocardial and 30% epicardial phenotypes, together with a 10 ms apico-basal action-potential-duration gradient[11]. Ventricular activation was initiated through an endocardial fast-activation layer mimicking the trifascicular conduction system and Purkinje–myocardial junctions[12]. Activation parameters were calibrated using beat-averaged 12-lead ECG biomarkers from a healthy female volunteer with myocardial mass comparable to the biventricular geometry, following established ECG-informed calibration strategies[13–15]. Standard 12-lead electrode positions were assigned according to anatomical landmark–based locations informed by established ECG-personalisation approaches, and pseudo-ECGs were computed at these locations for ECG calibration and biomarker extraction[13–15]. QT dispersion (QTd) was computed from the simulated 12-lead pseudo-ECGs as the difference between the longest and shortest measurable uncorrected QT intervals across leads. This measure was used alongside QRS duration and QTc to assess changes in inter-lead repolarisation variability across the staged HCM phenotypes and ionic-remodelling severities.

#### 1.3. Ventricular mechanics, loading, and electromechanical coupling

Ventricular mechanics were represented using a 3D dynamic hyperelastic active-stress finite-element formulation, governed by the balance of linear momentum in a total-Lagrangian framework and including inertial effects. The end-diastolic biventricular mesh was isotropically scaled down to define a diastasis reference configuration approximating a mechanically unloaded, near-zero-pressure state. Passive myocardial behaviour was modelled as a nearly incompressible, orthotropic hyperelastic material using the Holzapfel–Ogden constitutive law[16,17].

Physiological boundary conditions were applied to reproduce ventricular loading and pericardial constraint[18,19]. Time-varying pressure boundary conditions were prescribed independently on the left and right ventricular endocardial surfaces using a multiphase control strategy comprising diastolic filling, isovolumetric contraction, ejection, isovolumetric relaxation, and passive filling. During ejection, ventricular outflow was coupled to reduced-order systemic and pulmonary circulations using two-element Windkessel models. Pericardial interaction was represented by an elastic constraint of constant stiffness applied normal to the epicardial surface, limiting excessive expansion while preserving physiological deformation.

Electromechanical coupling was achieved by linking ToR-ORd calcium transients to active force generation through the Land myofilament formulation, forming the coupled ToR-ORd–Land framework[5,20]. The coupling was bidirectional, with intracellular calcium driving troponin binding and cross-bridge cycling, while troponin-bound calcium contributed to intracellular calcium buffering. Myocardial stretch and stretch rate influenced active-tension development, and deformation-dependent diffusion provided feedback from mechanics to electrical propagation. Active tension was incorporated into the mechanical equilibrium equations and superimposed on the passive stress response, enabling simulation of excitation–contraction coupling and its effects on ventricular deformation and pressure–volume behaviour.

#### 1.4. High-performance-computing configuration and simulation protocol

Simulations were run on ARCHER2 using 12 compute nodes, 128 MPI tasks per node, and one CPU core per task, corresponding to 1,536 MPI tasks/CPU cores per run. Each run simulated four cardiac cycles with a cycle length of 1 second. The first cycle was excluded from analysis to reduce the influence of initial conditions. Near-periodic behaviour was assessed by comparing the third and fourth cycles, which showed negligible differences; biomarkers were therefore extracted from the fourth and final simulated cycle.

##### 1.5. Model-dependent parameters

Key electrophysiological, ionic-remodelling, active-tension, geometric, haemodynamic, and mechanical parameters used to construct the healthy-control model and staged HCM phenotypes are summarised in Supplementary Table S1. Parameters were held fixed across simulations unless explicitly modified to represent a predefined disease mechanism or haemodynamic boundary condition. End-diastolic pressure and arterial resistance were adjusted across stages to represent haemodynamic changes associated with HCM progression. Elevated systemic vascular resistance has been reported in patients with non-obstructive HCM[21]. Prescribed end-diastolic pressure (PENDD) increased from 12,000 Ba (approximately 9.0 mmHg) in control to 14,000, 18,000, and 24,000 Ba (approximately 10.5, 13.5, and 18.0 mmHg) in Stages I–III, respectively. Arterial resistance ($R_{pres}$) increased from 300 Ba s $cm^{-3}$ in control to 350, 400, and 550 Ba s $cm^{-3}$ in Stages I–III, respectively. These parameters defined the haemodynamic environment of each disease stage. Passive myocardial stiffness was increased threefold only in Stage III, with the modified parameters highlighted in bold in Supplementary Table S1. Within each stage, ionic-remodelling comparisons were performed without changes to ventricular geometry, passive myocardial stiffness, or haemodynamic boundary conditions.

#### 2. Effects of graded ionic remodelling across progressive HCM stages

This section examines how graded secondary ionic remodelling modifies ventricular haemodynamics and simulated ECG morphology across the staged HCM phenotypes (Supplementary Figure S2; Supplementary Table S2). Across all stages, the mutation-only phenotype shifted the pressure–volume relationship towards smaller ventricular volumes and higher systolic pressures compared with the healthy control. The control simulation had an EDV of 142.7 mL, ESV of 56.5 mL, peak LV pressure of 106.3 mmHg, and EF of 60.4%. In the mutation-only simulations, EDV progressively decreased from 136.1 mL in Stage I to 132.9 mL in Stage II and 125.9 mL in Stage III, while peak LV pressure increased from 113.8 to 120.5 and 125.3 mmHg, respectively. EF remained preserved or mildly increased across the staged mutation-only phenotypes, reaching 64.3% in Stage I, 66.9% in Stage II, and 64.4% in Stage III.

Within each disease stage, increasing ionic-remodelling severity produced a further leftward displacement of the pressure–volume loop (Supplementary Figure S2). This was reflected by progressive reductions in EDV and ESV, most prominently under extreme ionic remodelling, where EDV decreased to 124.8 mL in Stage I, 121.3 mL in Stage II, and 113.9 mL in Stage III. Peak LV pressure was also attenuated with increasing ionic-remodelling severity, falling from 113.8 to 108.2 mmHg in Stage I, from 120.5 to 113.7 mmHg in Stage II, and from 125.3 to 116.9 mmHg in Stage III. Although EF remained preserved at 63.4%, 65.8%, and 62.9% under extreme ionic remodelling in Stages I–III, respectively, stroke volume decreased to 79.1, 79.8, and 71.6 mL. The reduction was greatest in Stage III, indicating that preserved EF did not imply preserved stroke volume in the smaller, mechanically constrained ventricle.

Electrophysiologically, graded ionic remodelling predominantly affected ventricular repolarisation. In lead V1, mutation-only remodelling produced modest QT prolongation compared with control, increasing QT interval from 350 ms to 364, 371, and 367 ms across Stages I–III. With increasing ionic-remodelling severity, repolarisation abnormalities became progressively more evident, with marked QT prolongation and T-wave changes across stages. Under extreme ionic remodelling, QT interval in V1 prolonged to 530, 555, and 558 ms in Stages I–III, respectively. QRS changes were smaller and less consistent than QT prolongation and were therefore not interpreted as the dominant electrophysiological effect. A similar pattern of severity-dependent repolarisation delay was observed in lead V5 (Supplementary Figure S2). Mutation-only remodelling produced only small increases in QT interval, from 351 ms in control to 356, 367, and 369 ms in Stages I–III. Graded ionic remodelling produced a stepwise prolongation of QT interval in V5, reaching 530 ms in Stage I, 543 ms in Stage II, and 556 ms in Stage III under the extreme ionic-remodelling condition. QT dispersion showed a similar overall increase with ionic-remodelling severity (Supplementary Table S2). Control QTd was 40.3 ± 5.5 ms, increasing to 42.3 ± 0.5, 48.1 ± 2.1, and 55.9 ± 2.0 ms in Stages I–III without ionic remodelling, respectively. With extreme ionic remodelling, QTd increased further to 73.4 ± 0.1, 78.4 ± 1.2, and 78.8 ± 0.5 ms in Stages I–III, respectively. The increase was generally severity dependent, although Stage III showed a slightly higher QTd under severe (79.1 ± 0.5 ms) than extreme (78.8 ± 0.5 ms) ionic remodelling. These findings indicate that secondary ionic remodelling increased inter-lead variation in QT duration in addition to prolonging the QT interval.

Together, these findings indicate that secondary ionic remodelling predominantly amplified repolarisation delay, QT dispersion, and T-wave abnormalities, while its haemodynamic effects were expressed mainly as reduced ventricular volumes, lower stroke volume at greater remodelling severity, and modest attenuation of peak systolic pressure.

**Supplementary Table S1.** Electrical and mechanical parameterisation of control and staged HCM models.

| Category | Parameter | Control | HCM | | |
|---|---|---|---|---|---|
| | | | Stage I | Stage II | Stage III |
| Diffusivity in fibre, sheet and sheet-normal directions | $d_f$ (cm/mS) | 0.001591 | 0.001591 | 0.001591 | 0.001591 |
| | $d_s$ (cm/mS) | 0.001038 | 0.001038 | 0.001038 | 0.001038 |
| | $d_n$ (cm/mS) | 0.00129 | 0.00129 | 0.00129 | 0.00129 |
| Compressibility | bulk modulus $K_{ct}$ (Ba) | 12185000 | 12185000 | 12185000 | 12185000 |
| Passive stiffness in isotropic matrix, fibre, sheet and fibre-sheet direction | a (Ba) | 100 | 100 | 100 | **300** |
| | b | 9.242 | 9.242 | 9.242 | 9.242 |
| | $a_f$ (Ba) | 500 | 500 | 500 | **1500** |
| | $b_f$ | 15.972 | 15.972 | 15.972 | 15.972 |
| | $a_s$ (Ba) | 200 | 200 | 200 | **600** |
| | $b_s$ | 10.446 | 10.446 | 10.446 | 10.446 |
| | $a_{fs}$ (Ba) | 200 | 200 | 200 | **600** |
| | $b_{fs}$ | 11.602 | 11.602 | 11.602 | 11.602 |
| Pressure at diastasis | PZERO (Ba) | 5000 | 5000 | 5000 | 5000 |
| Prescribed end-diastolic pressure | PENDD (Ba) | 12000 | **14000** | **18000** | **24000** |
| Pressure at end of isovolumetric relaxation | PPOST (Ba) | 10000 | 10000 | 10000 | 10000 |
| Aortic pressure | PART0 (Ba) | 90000 | 90000 | 90000 | 90000 |

| | | | | | |
|---|---|---|---|---|---|
| Arterial compliance | $C_{pres}$ ($cm^3$ $Ba^{-1}$) | 0.00055 | 0.00055 | 0.00055 | 0.00055 |
| Arterial resistance | $R_{pres}$ (Ba s $cm^{-3}$) | 300 | **350** | **400** | **550** |
| Pericardial stiffness | $K_{epi}$ | 20000 | 20000 | 20000 | 20000 |
| Penalty parameters for isovolumetric contraction | $C_v$ ($cm^3$ $s^{-1}$ $Ba^{-1}$) | 1 12 | 1 12 | 1 12 | 1 12 |
| Penalty parameters for isovolumetric relaxation | $C_v$ ($cm^3$ $s^{-1}$ $Ba^{-1}$) | 0.1 0.15 | 0.1 0.15 | 0.1 0.15 | 0.1 0.15 |
| Penalty parameters for passive filling | $C_p$, $C_v$ ($cm^3$ $Ba^{-1}$, $cm^3$ $s^{-1}$ $Ba^{-1}$) | 1 30 | 1 30 | 1 30 | 1 30 |

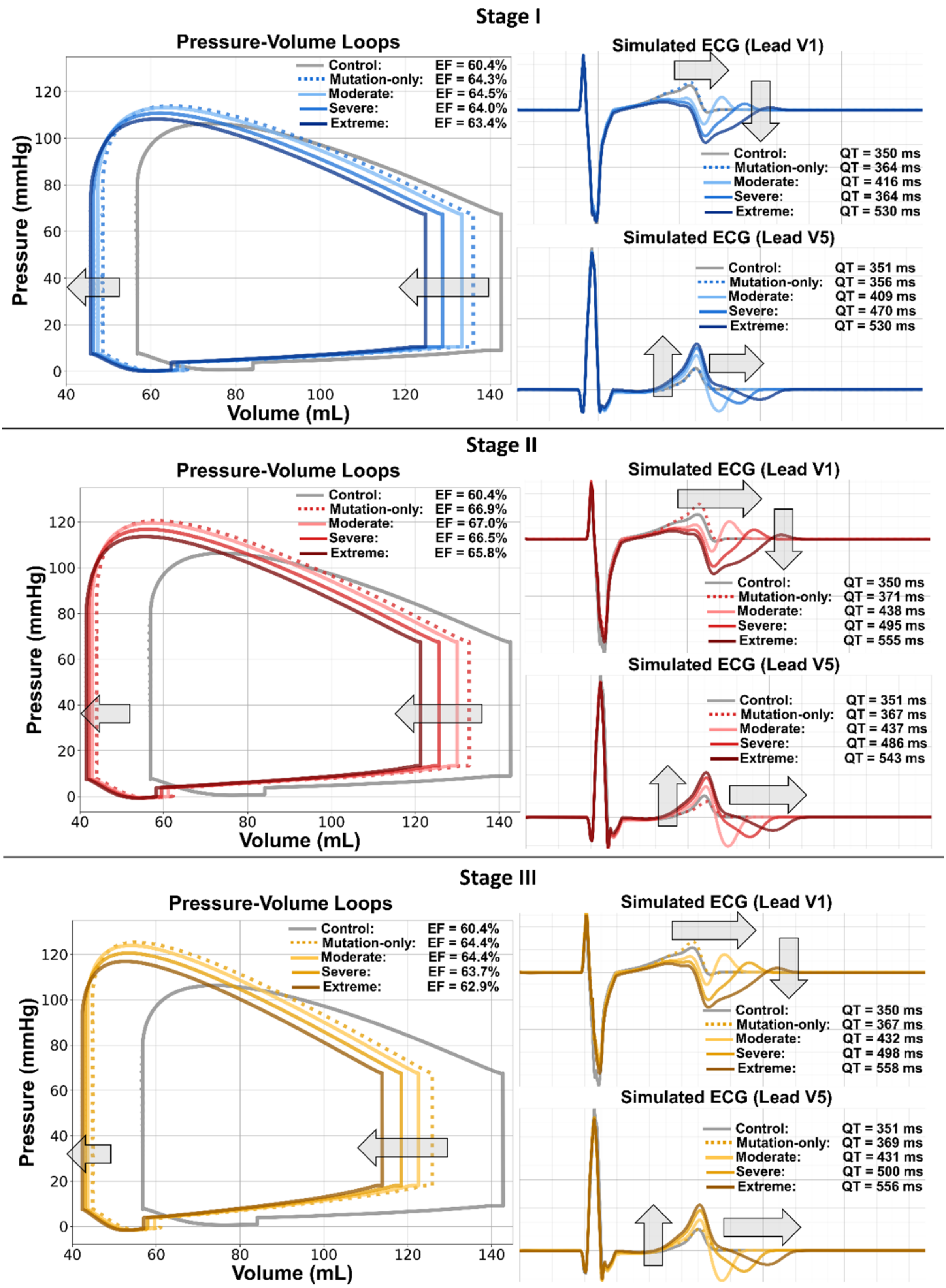


**Supplementary Figure S2. Left ventricular pressure–volume loops and simulated V1/V5 ECGs during staged HCM remodelling.** Stage I represents non-hypertrophic MYH7 disease, Stage II septal hypertrophy, and Stage III increased stiffness. Each stage includes mutation-only and moderate, severe, and extreme ionic-remodelling profiles. Control is shown in grey; mutation-only as dashed traces; and increasing ionic remodelling as progressively darker solid traces. EF and QT are reported for each condition.

**Supplementary Table S2.** Electrophysiological, haemodynamic, and deformation biomarkers across staged HCM progression and secondary ionic-remodelling profiles.

| Biomarker | Unit | Control | Stage | No ionic | Ionic Moderate | Ionic Severe | Ionic Extreme |
|---|---|---|---|---|---|---|---|
| **Electrophysiological biomarkers** | | | | | | | |
| QRSd | ms | 89.4 ± 9.7 | Stage I | 90.8 ± 10.7 | 88.8 ± 8.9 | 92.2 ± 5.6 | 98.6 ± 12.5 |
| | | | Stage II | 102.4 ± 4.8 | 102.0 ± 6.7 | 102.7 ± 10.6 | 109.7 ± 14.8 |
| | | | Stage III | 99.4 ± 7.1 | 102.2 ± 8.5 | 107.6 ± 12.0 | 112.5 ± 15.9 |
| QTc | ms | 354.7 ± 6.0 | Stage I | 361.5 ± 12.4 | 406.4 ± 13.9 | 457.2 ± 13.9 | 519.5 ± 22.4 |
| | | | Stage II | 371.1 ± 9.3 | 433.9 ± 6.0 | 486.5 ± 13.1 | 541.0 ± 18.5 |
| | | | Stage III | 372.4 ± 7.6 | 426.7 ± 6.6 | 490.6 ± 18.3 | 547.9 ± 22.8 |
| QTd | ms | 40.3 ± 5.5 | Stage I | 42.3 ± 0.5 | 61.3 ± 0.1 | 69.6 ± 5.0 | 73.4 ± 0.1 |
| | | | Stage II | 48.1 ± 2.1 | 66.8 ± 0.2 | 73.9 ± 0.5 | 78.4 ± 1.2 |
| | | | Stage III | 55.9 ± 2.0 | 72.1 ± 0.3 | 79.1 ± 0.5 | 78.8 ± 0.5 |
| **Haemodynamic biomarkers** | | | | | | | |
| EDV | mL | 142.7 | Stage I | 136.1 | 133.4 | 128.8 | 124.8 |
| | | | Stage II | 132.9 | 130.0 | 125.7 | 121.3 |
| | | | Stage III | 125.9 | 122.6 | 118.5 | 113.9 |
| ESV | mL | 56.5 | Stage I | 48.5 | 47.4 | 46.4 | 45.7 |
| | | | Stage II | 44.0 | 42.9 | 42.1 | 41.5 |
| | | | Stage III | 44.8 | 43.6 | 43.0 | 42.3 |
| SV | mL | 86.2 | Stage I | 87.6 | 86.0 | 82.4 | 79.1 |
| | | | Stage II | 88.9 | 87.1 | 83.6 | 79.8 |
| | | | Stage III | 81.1 | 79.0 | 75.5 | 71.6 |
| EF | % | 60.4 | Stage I | 64.3 | 64.5 | 64.0 | 63.4 |
| | | | Stage II | 66.9 | 67.0 | 66.5 | 65.8 |
| | | | Stage III | 64.4 | 64.4 | 63.7 | 62.9 |

**Supplementary Table S2 (continued).** Electrophysiological, haemodynamic, and deformation biomarkers across staged HCM progression and secondary ionic-remodelling profiles.

| Biomarker | Unit | Control | Stage | No ionic | Ionic Moderate | Ionic Severe | Ionic Extreme |
|---|---|---|---|---|---|---|---|
| **Haemodynamic biomarkers** | | | | | | | |
| EDP | mm Hg | 9.0 | Stage I | 10.5 | 10.5 | 10.5 | 10.5 |
| | | | Stage II | 13.5 | 13.5 | 13.5 | 13.5 |
| | | | Stage III | 18.0 | 18.0 | 18.0 | 18.0 |
| SP | mm Hg | 106.3 | Stage I | 113.8 | 113.1 | 110.7 | 108.2 |
| | | | Stage II | 120.5 | 119.6 | 116.8 | 113.7 |
| | | | Stage III | 125.3 | 123.9 | 120.5 | 116.9 |
| **Deformation biomarkers** | | | | | | | |
| GLS (Ell) | % | -17.9 ± 0.5 | Stage I | -16.6 ± 0.3 | -16.5 ± 0.2 | -16.1 ± 0.6 | -15.6 ± 1.2 |
| | | | Stage II | -17.2 ± 0.2 | -16.8 ± 0.1 | -15.7 ± 0.1 | -14.2 ± 0.1 |
| | | | Stage III | -16.0 ± 0.1 | -15.3 ± 0.1 | -13.9 ± 0.2 | -12.4 ± 0.1 |
| GRS (Err) | % | 54.8 ± 0.6 | Stage I | 54.0 ± 1.6 | 48.2 ± 2.0 | 40.9 ± 1.2 | 39.2 ± 1.1 |
| | | | Stage II | 52.3 ± 2.5 | 49.1 ± 2.4 | 42.6 ± 1.8 | 39.6 ± 1.0 |
| | | | Stage III | 51.7 ± 3.8 | 47.3 ± 3.8 | 41.4 ± 2.1 | 38.2 ± 1.1 |
| GCS (Ecc) | % | -18.8 ± 0.1 | Stage I | -21.0 ± 0.1 | -20.4 ± 0.3 | -18.7 ± 0.6 | -17.3 ± 0.6 |
| | | | Stage II | -23.0 ± 0.8 | -21.8 ± 0.7 | -18.9 ± 0.5 | -16.6 ± 0.2 |
| | | | Stage III | -25.2 ± 1.3 | -23.6 ± 1.3 | -20.1 ± 1.1 | -17.4 ± 0.4 |

### 3. Strain biomarkers identify stage-dependent mechanical deterioration in HCM

This section evaluates how the staged HCM phenotypes and graded secondary ionic remodelling altered median septal strain over the normalised cardiac cycle (Supplementary Figure S3).

Simulated myocardial strain was obtained as Green–Lagrange strain referenced to the diastasis configuration. The strain tensor was projected onto the local circumferential, radial, and longitudinal directions to obtain the corresponding directional components:

$$E_\alpha(t) = \mathbf{e}_\alpha^{\mathrm{T}}\, \mathbf{E}_{\mathrm{sim}}(t)\, \mathbf{e}_\alpha, \qquad \alpha \in \{r, c, l\}$$

where $\mathbf{E}_{\mathrm{sim}}(t)$ is the Green–Lagrange strain tensor and $\mathbf{e}_\alpha$ is the corresponding unit direction. To express strain relative to end-diastole (ED) and allow comparison with engineering-strain measurements, the directional components were converted according to:

$$\varepsilon_\alpha^{\mathrm{ED}}(t) = \sqrt{\frac{2E_\alpha(t) + 1}{2E_\alpha(t_{\mathrm{ED}}) + 1}} - 1$$

where $E_\alpha(t_{\mathrm{ED}})$ denotes the Green–Lagrange strain at end-diastole relative to diastasis. Median strain curves were calculated from the selected septal nodes, with circumferential and radial components evaluated in the short-axis region and longitudinal strain in the long-axis region. Strain values are reported as percentages.

Across stages, the simulations showed reduced radial strain and longitudinal shortening, accompanied by progressive attenuation of late-diastolic strain recovery. These effects were most pronounced with increasing ionic-remodelling severity and in the more advanced HCM stages.

In the control simulation, septal deformation was characterised by peak circumferential strain of −16.8%, radial strain of 72.2%, and longitudinal strain of −20.2% (Supplementary Figure S3). Stage I mutation-only remodelling increased peak circumferential shortening to −20.1%, while peak radial strain remained near the control value at 70.3%. In contrast, longitudinal shortening was already reduced to −17.8%, indicating early impairment of longitudinal mechanics despite preserved or enhanced circumferential deformation. With increasing ionic-remodelling severity, radial and longitudinal strain declined progressively. Under extreme ionic remodelling in Stage I, radial strain fell to 38.9% and longitudinal strain to −12.6%, while circumferential strain remained relatively preserved at −18.7%. These findings suggest that early ionic remodelling affected radial and longitudinal deformation more strongly than circumferential shortening.

In Stage II, septal hypertrophic remodelling produced a marked increase in circumferential shortening in the mutation-only condition, with peak Ecc reaching −26.4%. However, this was accompanied by a substantial reduction in radial strain to 44.7% and longitudinal shortening to −16.0%, consistent with a redistribution of septal mechanics rather than uniform preservation of contractile function. Increasing ionic-remodelling severity progressively blunted this deformation pattern. Peak circumferential strain decreased from −26.4% in the mutation-only condition to −15.0% under extreme ionic remodelling, radial strain declined from 44.7% to 31.8%, and longitudinal strain decreased from −16.0% to −12.8%. Thus, in the hypertrophic stage, ionic remodelling attenuated circumferential and longitudinal shortening and radial strain.

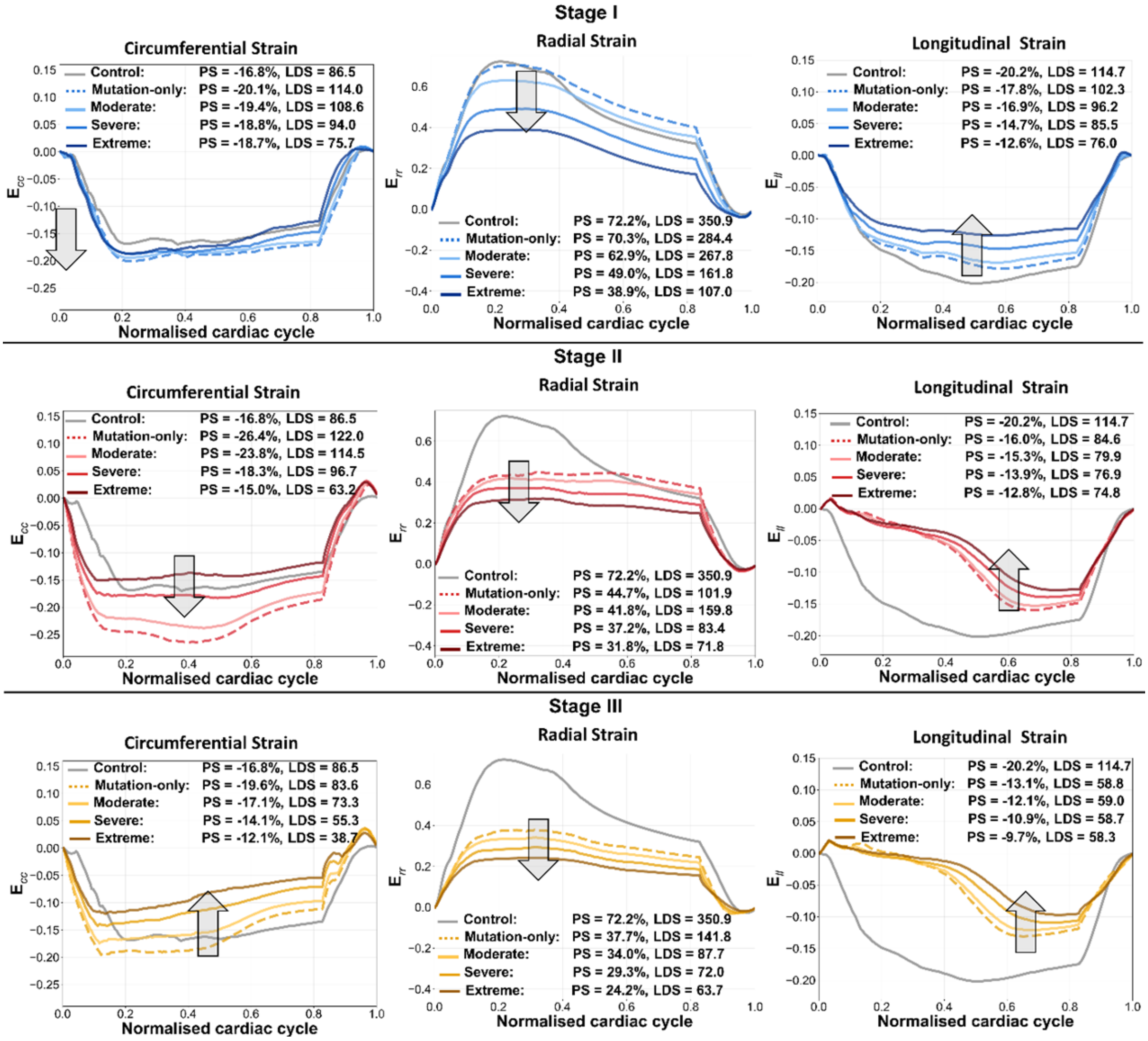


**Supplementary Figure S3.** Median septal strain across staged HCM phenotypes and graded ionic remodelling. Median septal circumferential (Ecc), radial (Err), and longitudinal (Ell) strain over the normalised cardiac cycle are shown for the healthy control and Stage I, Stage II, and Stage III HCM phenotypes. The grey trace denotes the healthy-control reference; dashed traces denote mutation-only simulations; and progressively darker solid traces denote moderate, severe, and extreme ionic-remodelling profiles. Peak strain (PS) and late-diastolic slope (LDS) are annotated for each condition. Arrows indicate the dominant direction of change with increasing ionic-remodelling severity.

Stage III demonstrated the most advanced impairment of septal mechanics (Supplementary Figure S3). Compared with control, all Stage III profiles showed reduced radial and longitudinal deformation, with the greatest reductions under extreme ionic remodelling. Peak radial strain decreased from 72.2% in control to 37.7% in the Stage III mutation-only condition and to 24.2% with extreme ionic remodelling. Longitudinal strain similarly declined from −20.2% in control to −13.1% in mutation-only Stage III and to −9.7% under extreme ionic remodelling. Circumferential strain also became progressively attenuated, falling from −19.6% in Stage III mutation-only to −12.1% with extreme ionic remodelling. These changes were consistent with reduced septal deformation across all three principal strain directions in the stiffened Stage III phenotype.

Late-diastolic strain recovery was also progressively impaired across the staged phenotypes and with increasing ionic-remodelling severity. For circumferential strain, late-diastolic slope increased in mutation-only Stage I and Stage II relative to control, but declined substantially with increasing ionic-remodelling severity, reaching 75.7%/cycle in Stage I extreme, 63.2%/cycle in Stage II extreme, and 38.7%/cycle in Stage III extreme. Radial strain recovery showed a similar deterioration, with late-diastolic slope decreasing from 350.9%/cycle in control to 107.0%/cycle in Stage I extreme, 71.8%/cycle in Stage II extreme, and 63.7%/cycle in Stage III extreme. Longitudinal late-diastolic recovery also declined progressively, from 114.7%/cycle in control to 76.0, 74.8, and 58.3%/cycle under extreme ionic remodelling in Stages I–III, respectively.

Together, these results show that the staged HCM phenotypes exhibited distinct alterations in septal mechanics, while graded ionic remodelling amplified mechanical dysfunction across the cardiac cycle. The dominant effects were reduced radial strain, impaired longitudinal shortening, and delayed or attenuated diastolic strain recovery. Circumferential strain was initially preserved or increased, particularly in Stage II, but became progressively blunted with increasing ionic-remodelling severity and in the stiffened Stage III phenotype.

### 4. Spatiotemporal electromechanical remodelling across progressive HCM stages

This section examines the spatiotemporal evolution of transmembrane potential and ventricular deformation across staged HCM phenotypes at four representative phases of the cardiac cycle: end-diastole, QRS, ejection, and end-systole (Supplementary Figure S4). For simulations labelled as ionic remodelling, the extreme ionic-remodelling profile is shown. Across all cases, end-diastole was characterised by near-resting transmembrane potential and negligible displacement relative to the end-diastolic reference configuration. The membrane-potential maps showed broadly similar resting patterns across the staged phenotypes.

During QRS, all phenotypes demonstrated rapid ventricular depolarisation with limited mechanical displacement, consistent with electrical activation preceding substantial deformation. The spatial distribution of depolarisation was broadly comparable across the staged phenotypes. Local displacement during QRS remained small across all cases, ranging from 1.9 to 3.0 mm.

During ejection, ventricular deformation increased substantially across all phenotypes, with differences in mechanical displacement between ionic and non-ionic conditions. The accompanying changes in displacement were stage dependent: local displacement was lower with extreme ionic remodelling in Stage I (4.1 vs 6.3 mm without ionic remodelling), Stage II (6.0 vs 6.5 mm), and Stage III (6.2 vs 8.2 mm), showing that the mechanical response associated with ionic remodelling differed between the non-hypertrophic and structurally remodelled phenotypes.

At end-systole, local transmembrane potential returned towards resting values in all cases, but residual deformation differed across stages. The control simulation showed local displacement of 8.3 mm. Non-ionic HCM remodelling showed stage-dependent differences in local end-systolic displacement relative to control, reaching 6.9 mm in Stage I, 8.9 mm in Stage II, and 12.0 mm in Stage III. With extreme ionic remodelling, end-systolic displacement was reduced in Stage I (5.0 vs 6.9 mm without ionic remodelling), Stage II (6.9 vs 8.9 mm), and Stage III (8.7 vs 12.0 mm). These results show that extreme ionic remodelling was associated with reduced local ventricular displacement across all three HCM stages, with the largest absolute reduction observed in the stiffened Stage III phenotype.

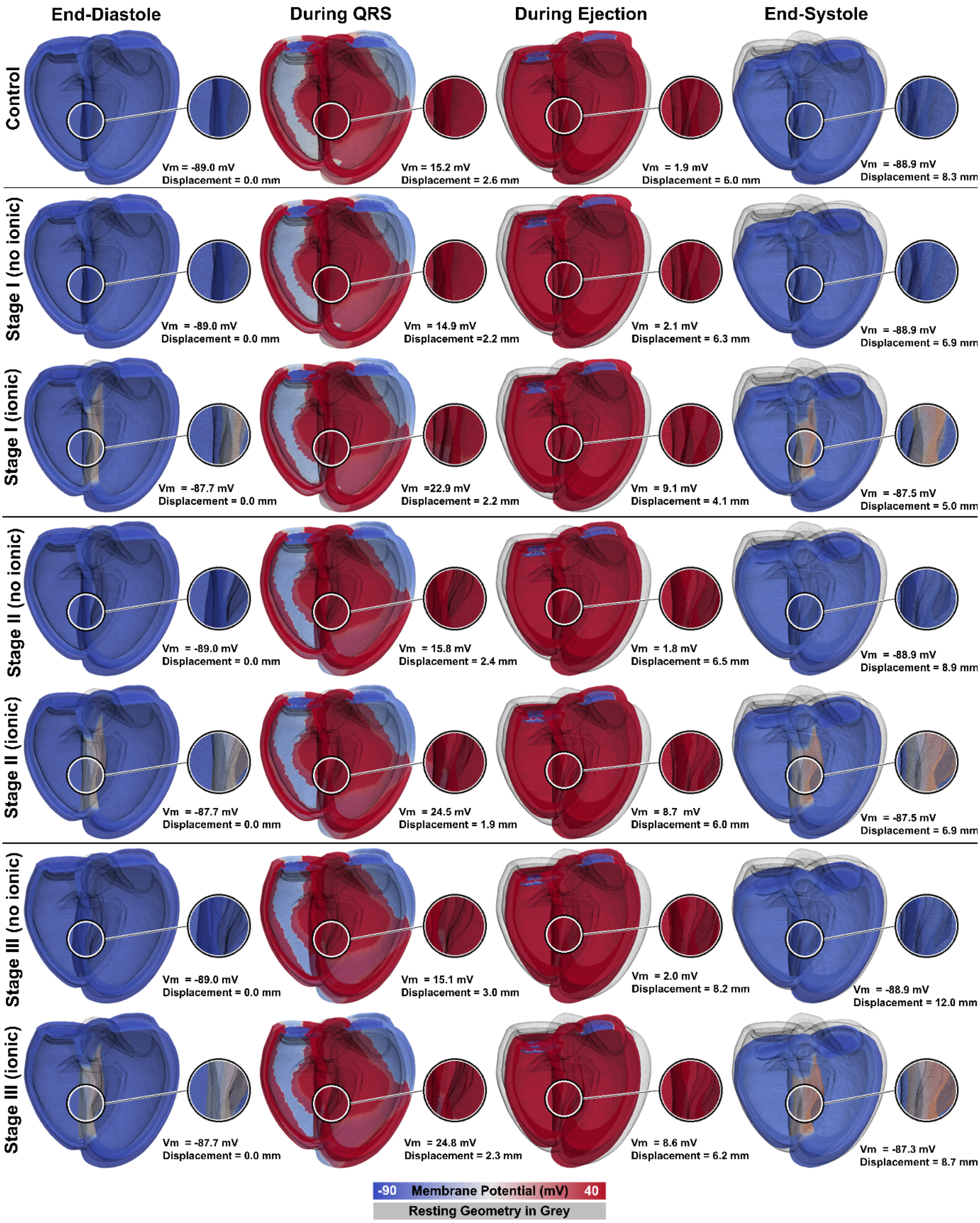


**Supplementary Figure S4**. Spatiotemporal evolution of transmembrane potential and ventricular deformation across staged HCM phenotypes. Rows show the healthy control and staged HCM phenotypes without ionic remodelling and with extreme ionic remodelling. Columns show four representative phases of the cardiac cycle: end-diastole, QRS, ejection, and end-systole. The ventricular geometry is coloured by transmembrane potential, with the resting reference geometry overlaid in grey. Insets highlight a representative septal region. Annotations report local transmembrane potential (Vm) and the magnitude of displacement relative to end-diastole at a selected septal proxy location. The membrane-potential colour scale is fixed between −90 and +40 mV across all simulations, with values outside this range saturated. The figure illustrates phase-dependent electrical activation and repolarisation, together with stage-dependent reductions in local ventricular displacement associated with extreme ionic remodelling, particularly in the hypertrophic and stiffened phenotypes.
